# Molecular mapping of urinary complement peptides in kidney diseases

**DOI:** 10.1101/2021.06.24.21259458

**Authors:** Ralph Wendt, Justyna Siwy, Tianlin He, Agnieszka Latosinska, Thorsten Wiech, Peter F. Zipfel, Aggeliki Tserga, Antonia Vlahou, Lorenzo Catanese, Harald Rupprecht, Harald Mischak, Joachim Beige

**Author notes:** Correspondence: Joachim Beige, Professor of Medicine, Kuratorium for Dialysis and Transplantation / Dept. Nephrology, Hospital St. Georg, Martin-Luther-University Halle/Wittenberg, Delitzscher Str. 141 (Haus 54), D-04129 Leipzig. These authors contributed equally (Ralph Wendt, Justyna Siwy, Joachim Beige).

## Abstract

Defective complement activation has been associated with various types of kidney disease. This led to the hypothesis that specific urine complement fragments may be associated with kidney disease etiologies, and disease progression may be reflected by changes in these complement fragments. We investigated the occurrence of complement fragments in urine, their association with kidney function, proteinuria and disease etiology. Mass spectrometry based peptidomics data from the Human Urinary Proteome/Peptidome Database were extracted and the distribution of complement peptides in the different kidney disease etiologies and controls was investigated. All datasets with informations on disease/health status and detectable complement peptides were included (n=16027). Twenty-three different urinary peptides derived from complement proteins could be identified, originating from the complement proteins C3, C4 and complement factor B. For most C3-derived peptides an inverse association with eGFR was observed, while the majority of peptides derived from CFB demonstrated positive association with eGFR. Highest levels of significant C3 excretion relative to controls were seen in minimal change disease (MCD), focal segmental glomerulosclerosis (FSGS), membranous glomerulonephritis (MGN), lupus nephritis (LN), diabetic kidney disease (DKD), IgAN, membranoproliferative glomerulonephritis (MPGN), and C3-glomerulonephritis. In conclusion, several peptides derived from the complement proteins C3, C4 and factor B are significantly associated with specific kidney disease etiologies. These peptides may depict disease-specific complement activation, as well as damage to the glomerular basement membrane. Further targeted investigation of these peptides may provide new insight into disease pathophysiology and could possibly guide therapeutic decisions, especially when targeting complement factors.

## Introduction

The urinary proteome holds significant information on disease and disease pathophysiology [1]. The complement system is a crucial effector of the innate immune system and also involved in mechanisms of the adaptive immune system. It is activated via 3 different pathways and comprised of over 30 different plasma or membrane-bound proteins [2;3]. The complement system is increasingly recognized as an important mediator in the context of kidney diseases [4]. The kidney seems particularly susceptible to complement-mediated injury. Immune complexes are strong activators of the classical pathway of complement and are involved in various forms of glomerulonephritis, e.g. lupus nephritis (LN). Dysregulation of the alternative pathway of complement activation may lead to atypical hemolytic uremic syndrome (aHUS) and C3-glomerulonephritis (C3-GN) [5]. All complement activation pathways converge at the level of C3-convertase. Complement activation as well as modulation of complement activity is exerted on the tissue or cell surfaces, or in the circulation. Most studies of complement association with disease involve biomarkers of complement functions or complement factors in serum or plasma of their deposition on the tissue surface. Studies on complement excretion in urine are scarce and might add important non-interventional information on complement status and function in different diseases.

The availability of a comprehensive database of urine peptide and low molecular weight protein data [6] has generated the opportunity to perform in-depth analysis of urine peptides and low molecular weight proteins, based on large numbers of patient-derived datasets. Investigations have been performed either in the context of specific diseases or physiology (e.g. heart failure or obesity [7;8], or specific peptides (e.g. thymosin beta-4 or polymeric immunoglobulin receptor [9;10]). Given the limited data available on the complement fragments in urine and their association with kidney disease, a hypothesis was generated that complement-derived peptides in urine allow to monitor complement activation or a damage of the glomerular basement membrane, and may be an indirect measure of the complement activation. Therefore, we investigated the urinary peptidome for the presence of complement-derived fragments and in a next step assessed the distribution of these peptides in different chronic kidney disease (CKD) etiologies, to identify potential specific association of complement fragments with CKD etiologies.

## Methods

### Proteomic investigation, data retrieval and diagnosis assessment

For this study urinary proteome data obtained by capillary electrophoresis coupled to mass spectrometry (CE-MS) and stored in the Human Urine Proteome Database [11] were assessed. The CE-MS technology applied has been described in detail including reproducibility, repeatability, procedures for sample preparation, data evaluation and normalization [12]. This database currently contains urinary proteome/peptidome information on >80.000 samples, assessing a dataspace comprised in total of >100.000 peptides and low molecular weight proteins [6], as well as information on the peptide sequencing from LC-MS/MS and CE-MS/MS analysis, and anonymized clinical information of participants enrolled in the studies. More detailed information on the peptide sequencing was described elsewhere [13]. Underlying diagnosis related to histopathology findings, e.g. IgA nephropathy (IgAN) were obtained by kidney biopsy. If biopsy was not indicated as for instance in the majority of cases with acute kidney failure and kidney stones, diagnoses of such scenarios were obtained by assessment of the available clinical information. All underlying studies were conducted to conform to regulations on the protection of individuals participating in medical research and in accordance with the principles of the Declaration of Helsinki (2013) and had received ethical approval by the responsible institutional review boards. Written informed consent was obtained from all participants at the time of sampling. All data sets received were anonymized. The study was approved by the ethics committee of the Hannover Medical School, Germany (no. 3116-2016). All proteomics datasets with the information about disease condition where at least one complement peptide could be detected were extracted. All datasets from subjects with disease other than kidney disease were excluded.

### Assessment of eGFR and statistics

Estimated glomerular filtration rate (eGFR) was calculated from serum creatinine using the Chronic Kidney Disease Epidemiology Collaboration (CKD-EPI) equation [14]. The association of proteomic data for all detectable complement fragments together with clinical data were investigated using a SPSS 22.0 software (IBM). Correlation analysis was performed based on log transformed values using the Spearman’s rank correlation coefficient.

### Correction for proteinuria

To correct for proteinuria, in a first step the abundance levels of all peptides originating from the same protein were combined into one variable per complement factor, “combined abundance”. This approach can further be justified based on the fact that most peptides observed are from the same region of the respective complement protein, hence are mutually exclusive (only one specific peptide can be generated from one complement molecule). The data were log transformed and correlation of the combined abundance with log proteinuria was assessed. From the regression equation ([peptide abundance] = a + b*[proteinuria] the value of b was determined and applied for each peptide as follows: [log peptide abundance corrected] = [log peptide abundance observed] - b*[log proteinuria].

## Results

All available sequencing data from Human Urinary Proteome/ Peptidome Database were investigated. A total of 23 complement fragments could be identified with high confidence. Tandem mass spectra from all 23 peptides are shown in **Supplementary Figure 1**. These urinary complement peptide fragments are presented in **Table 1**.

**Table 1:**
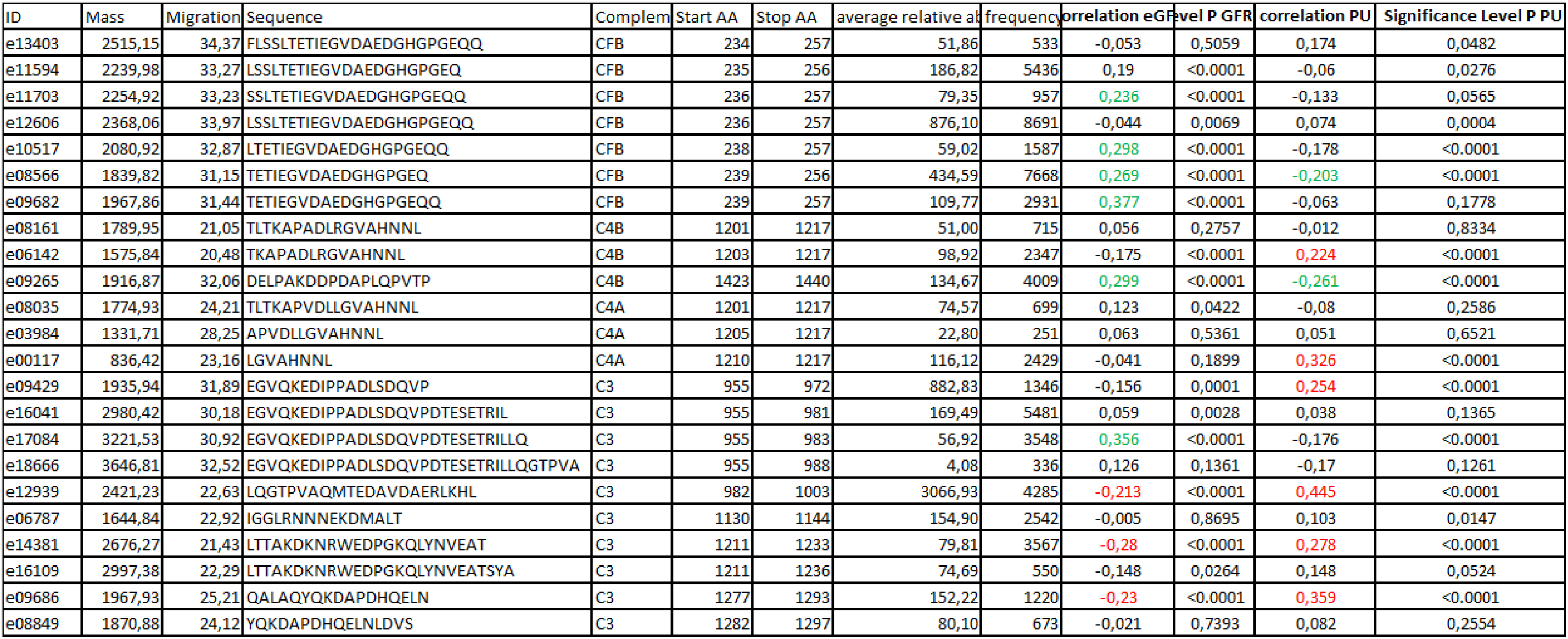
List of detected complement fragments. Given are an identification number (ID), peptide mass [Da], migration time [Min], amino acid sequence, parental complement protein, peptide position in the protein sequence, and average relative abundance and frequency of these peptides calculated based on the full dataset of 16027 individuals, along with correlation coefficient (rho) with eGFR and proteinuria (PU) of complement fragments together with the respective p-value.

All CE-MS datasets with available anonymized information on disease and with at least one of the complement peptides detectable were extracted from the database. This resulted in 16027 anonymized datasets that could be retrieved and employed in further analysis. A list of the number of subjects per disease etiology/condition is presented in **Table 2**.

**Table 2.** Distribution conditions/ etiologies for urinary peptidomics datasets included in the study.

| disease/condition | N |
| --- | --- |
| Acute kidney injury | 422 |
| ADPKD | 273 |
| Amyloidosis | 8 |
| Atypical hemolytic uremic syndrome | 8 |
| C3 glomerulopathy | 24 |
| CAKUT | 567 |
| Diabetes Mellitus | 4428 |
| Diabetic kidney disease | 1401 |
| Fanconi syndrom | 12 |
| FSGS | 126 |
| IgAN | 811 |
| MCD | 50 |
| MGN | 113 |
| Morbus Fabry | 66 |
| MPGN | 50 |
| Hypertensive nephrosclerosis | 104 |
| Nephrotic syndrom | 127 |
| NTx | 2300 |
| SLE | 20 |
| LN | 94 |
| Vaskulitis | 159 |
| Healthy control | 4864 |
|  | **16027** |

In the first step, potential correlation of complement fragments with eGFR was investigated. Data on eGFR were available from 8388 datasets. The results are presented in **Table 1**. Most complement C3 peptide fragments demonstrate a weak, yet significant inverse association with eGFR. In contrast, correlation of the complement factor B (CFB) fragments with eGFR was inconsistent: both, weak inverse and direct association were observed. As examples, the association of the most abundant peptide in urine from complement C3, L_982_QGTPVAQMTEDAVDAERLKHL_1003_ (e12939), and from CFB, L_235_SSLTETIEGVDAEDGHGPGEQ_257_ (e11594) with eGFR is shown in **Figure 1 A** and **C**. Most of the fragments from complement 4 A and B did not show association with eGFR. The urinary excretion of all detected complement fragments stratified by disease etiology/ condition is shown in **Figure 2**. The underlying data are presented in **Supplementary Table 1**. Shown are individual peptide excretion levels normalized to healthy controls. This was calculated by division of mean abundances per disease/condition with the mean corresponding abundance level in healthy controls. Highest relative levels (in reference to healthy controls) for LQGTPVAQMTEDAVDAERLKHL (e12939, complement C3) excretion were seen in minimal change disease (MCD), followed by nephrotic syndrome (NS), focal segmental glomerulosclerosis (FSGS), membranous glomerulonephritis (MGN), lupus nephritis (LN), diabetic kidney disease (DKD), IgAN, membranoproliferative glomerulonephritis (MPGN), and C3-glomerulonephritis (C3-GN), (in the order of median relative abundance).

**Figure 1.**
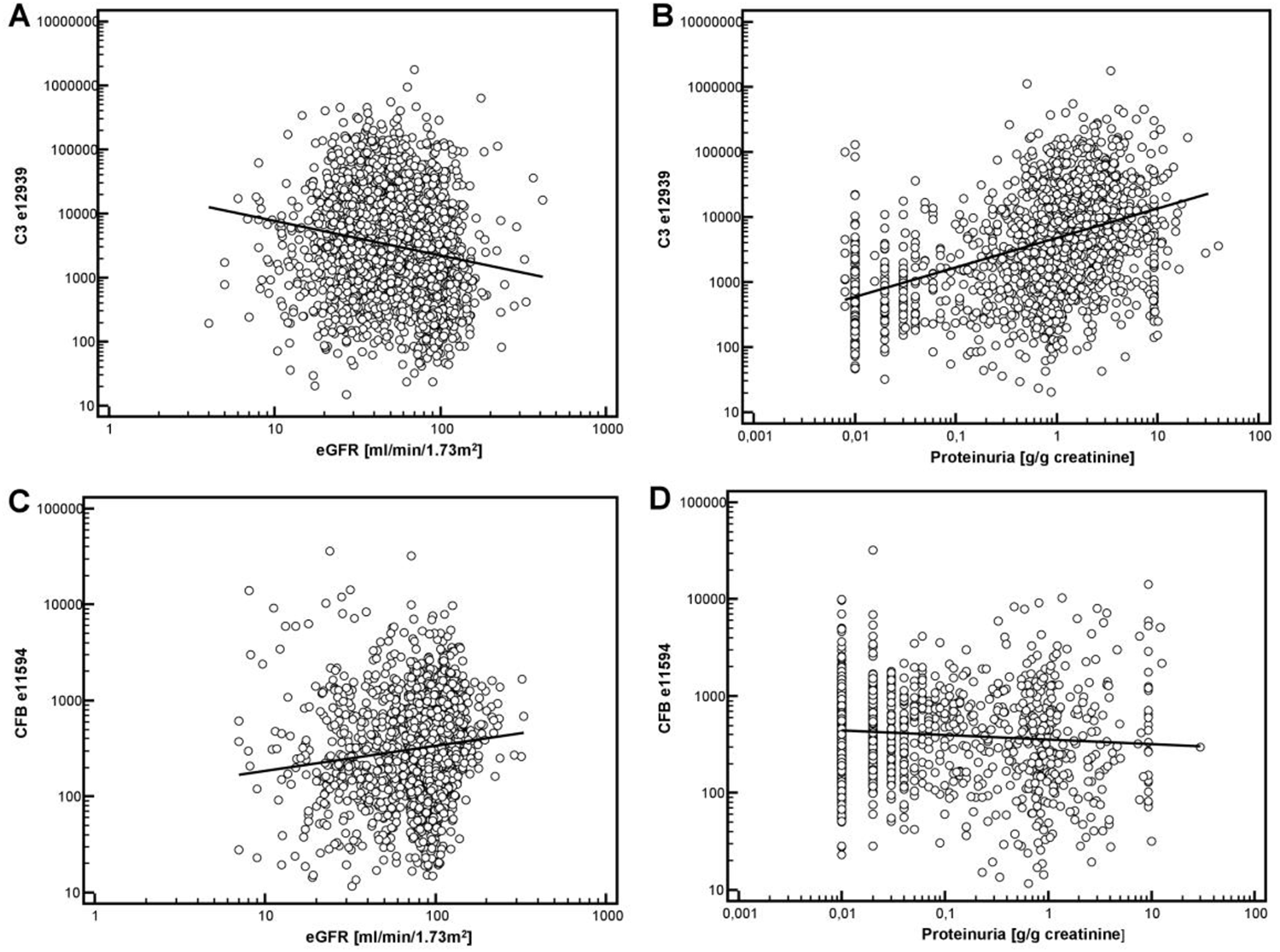
Association of complement derived peptides with eGFR and proteinuria. A: association of the most abundant peptide in urine from complement C3, L_982_QGTPVAQMTEDAVDAERLKHL_1003_ (e12939) with eGFR. B: association of e12939 with proteinuria. C: Association of L_235_SSLTETIEGVDAEDGHGPGEQ_257_ (e11594), from Complement factor B with eGFR. D: association of e11594 with proteinuria.

**Figure. 2:**
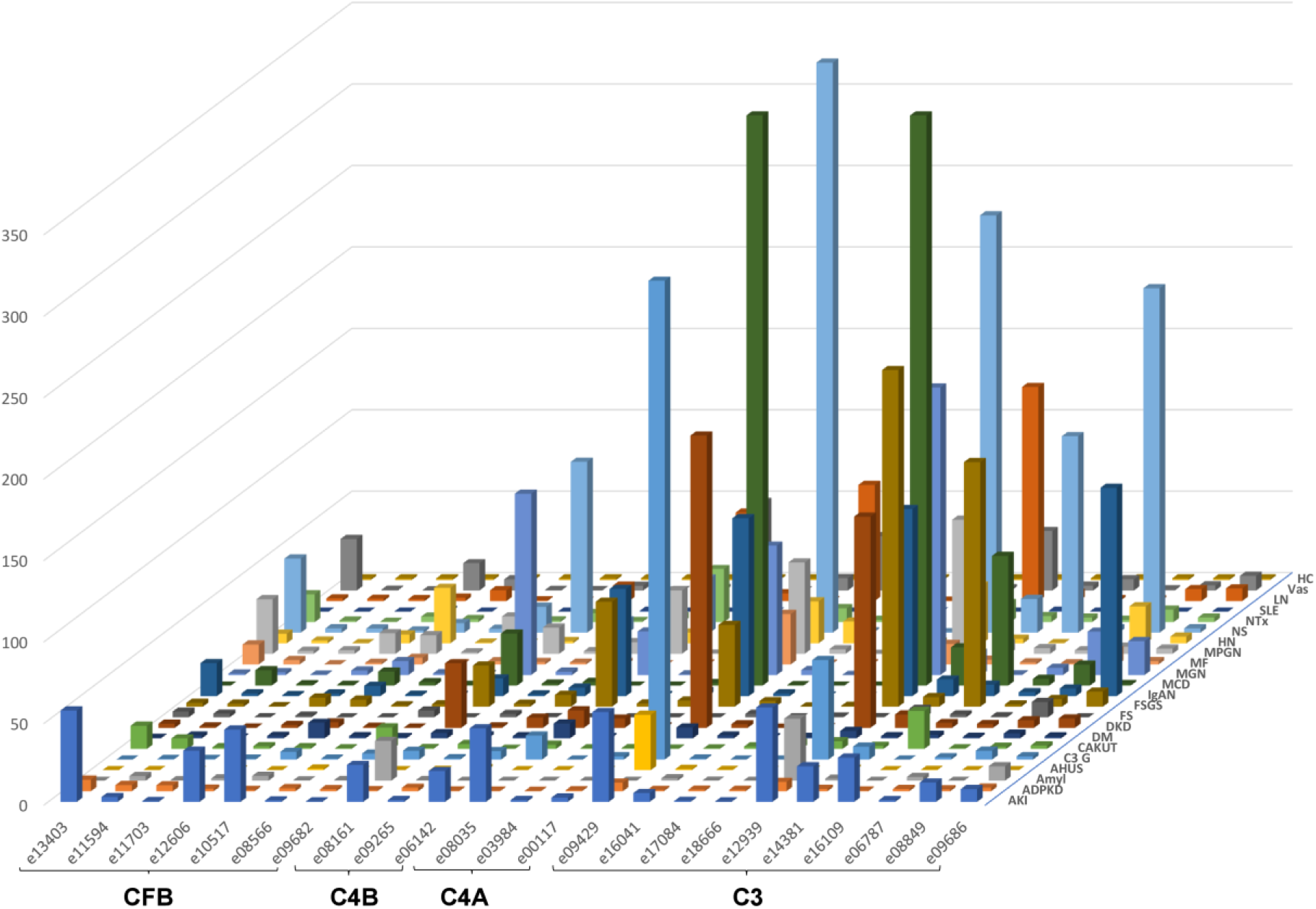
Peptide abundances of all complement fragments per disease or condition, normalized by median abundance in healthy controls. The ID of the peptides is given as indicated in Table 1. Peptides are sorted according to their origin, first peptides from CFB, then C4A and C4B, followed by C3. Within each protein, peptides are sorted based on the position of first amino acid.

Due to the high levels of excretion of complement C3-derived peptides in IgAN and the availability of a substantial number of follow-up data, we investigated the corresponding representative peptide in the independent PersTIgAN cohort [15]. In this cohort of 209 patients, LQGTPVAQMTEDAVDAERLKHL (e12939, complement C3) was significantly increased in the tertile of patients with the highest eGFR loss during follow-up, in comparison to the patients with the lowest eGFR loss (p=0.0225).

Highest median level of relative-to-healthy controls for LSSLTETIEGVDAEDGHGPGEQQ (e12606) excretion, a fragment of CFB, were seen in acute kidney injury (AKI), vasculitis, MPGN, and FSGS, (in the order of median relative abundance) with wide abundance distribution.

The above described inverse correlation between eGFR and C3 peptides may in fact not be direct, but due to different levels of proteinuria. In the next step we therefore investigated association of the abundance of complement peptides with proteinuria. Data on proteinuria were available from 4086 datasets. Examples on the association of peptide abundance and proteinuria for the above mentioned two peptides (e12939, e11594) are provided on **Figure 1B** and **D**. As shown in **Figure 1B**, a significant and relevant (rho=0.44) direct association of complement C3 peptide (e12939) with proteinuria could be detected. This association could in general also be detected when investigating the other complement C3 peptides (**Table 1**).

These observed associations of proteinuria with especially the complement C3 fragments indicate that the abundance of complement fragments in urine is in part the result of glomerular filtration of the respective peptide, and in part the result of proteinuria. When investigating the sequences of the observed complement peptides, it became evident that these originate in general from one specific region, with substantial overlap (see amino acid positions in **Table 1**). As such, these peptides are mutually exclusive, only one of these peptides can be generated from a single complement molecule. Based on this observation, the relative abundance values of the overlapping peptides were combined (added) into a single variable, per complement protein. The results of this approach, the distribution of the combined peptide abundances per complement component are shown in **Figure 3 A**, limited to those datasets where proteinuria values were available. As expected, a strong correlation was observed for the combined complement C3 peptides with proteinuria. A weak, yet significant correlation was also detected when investigating complement C4A and B, and a weak, inverse correlation could be detected for CFB (shown in **Supplementary Figure 2**). This “combined abundance” was used to correct for proteinuria, also to avoid overfitting when correction would be based on each single peptide. When applying this correction for the respective combined abundance levels, the correlation with proteinuria was lost (also shown in **Supplementary Figure 2)**. Subsequently, this correction for proteinuria calculated based on the change in “combined abundance” was applied for each peptide from each patient, and the distribution in the subjects where proteinuria was available was investigated in the different CKD etiology/ condition, with and without correction for proteinuria. The results are presented in **Figure 4**. In case of C3 fragments after adjustment for proteinuria, highest excretion was seen in MCD, C3-GN, DKD, FSGS, MGN, LN and also vasculitis. For CFB fragments corrected for proteinuria, high excretion was seen e.g. in AKI and MPGN.

**Figure. 3:**
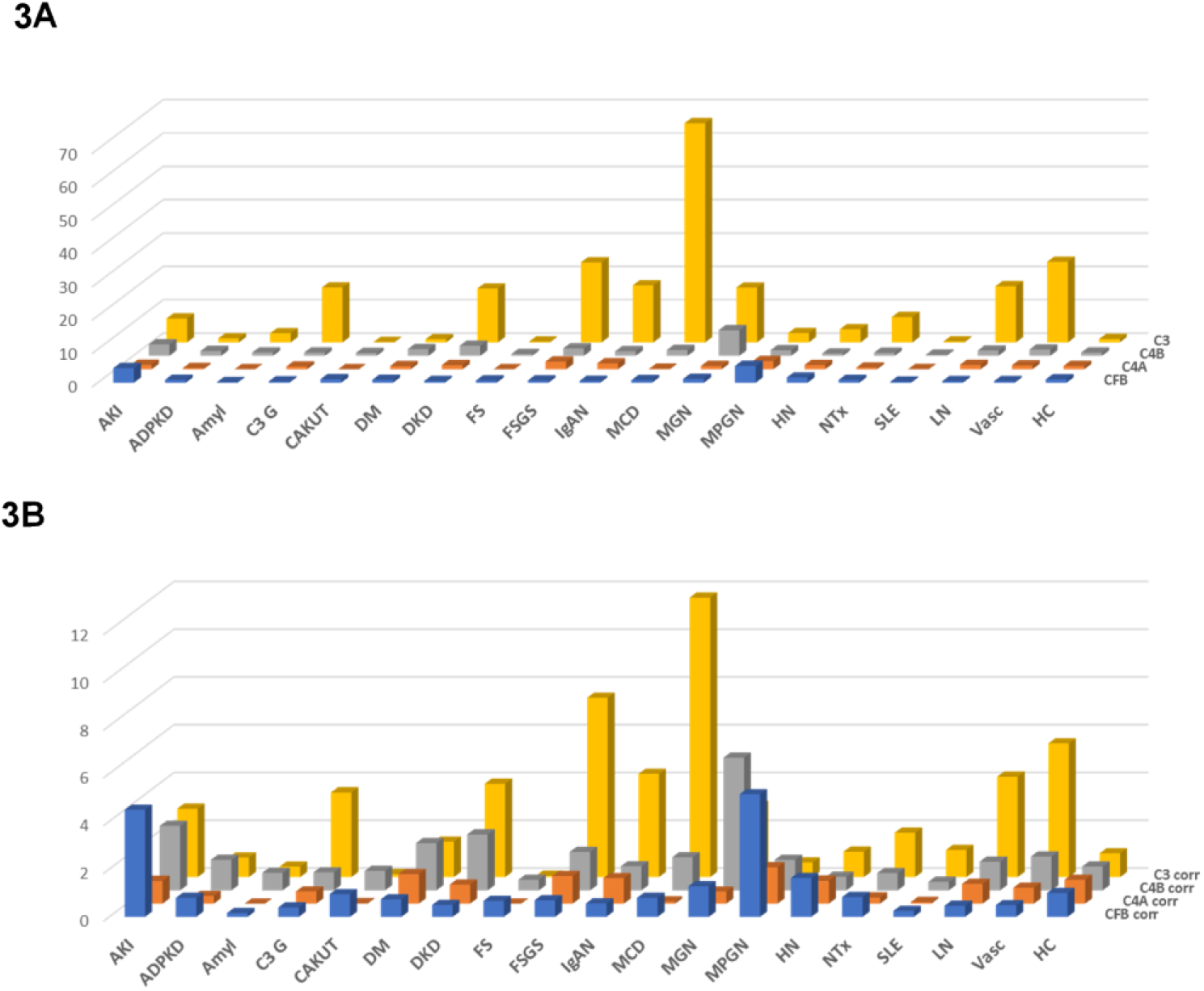
Relative abundances (normalized to the medium abundance in healthy controls) of the combined complement fragments from the four different members of the complement family, unadjusted or adjusted for proteinuria. A) Combined complement factor peptide abundances, unadjusted. B) Combined complement factor peptide abundances, adjusted for proteinuria. While adjustment for proteinuria induces some changes, the overall distribution is not substantially affected.

**Figure. 4:**
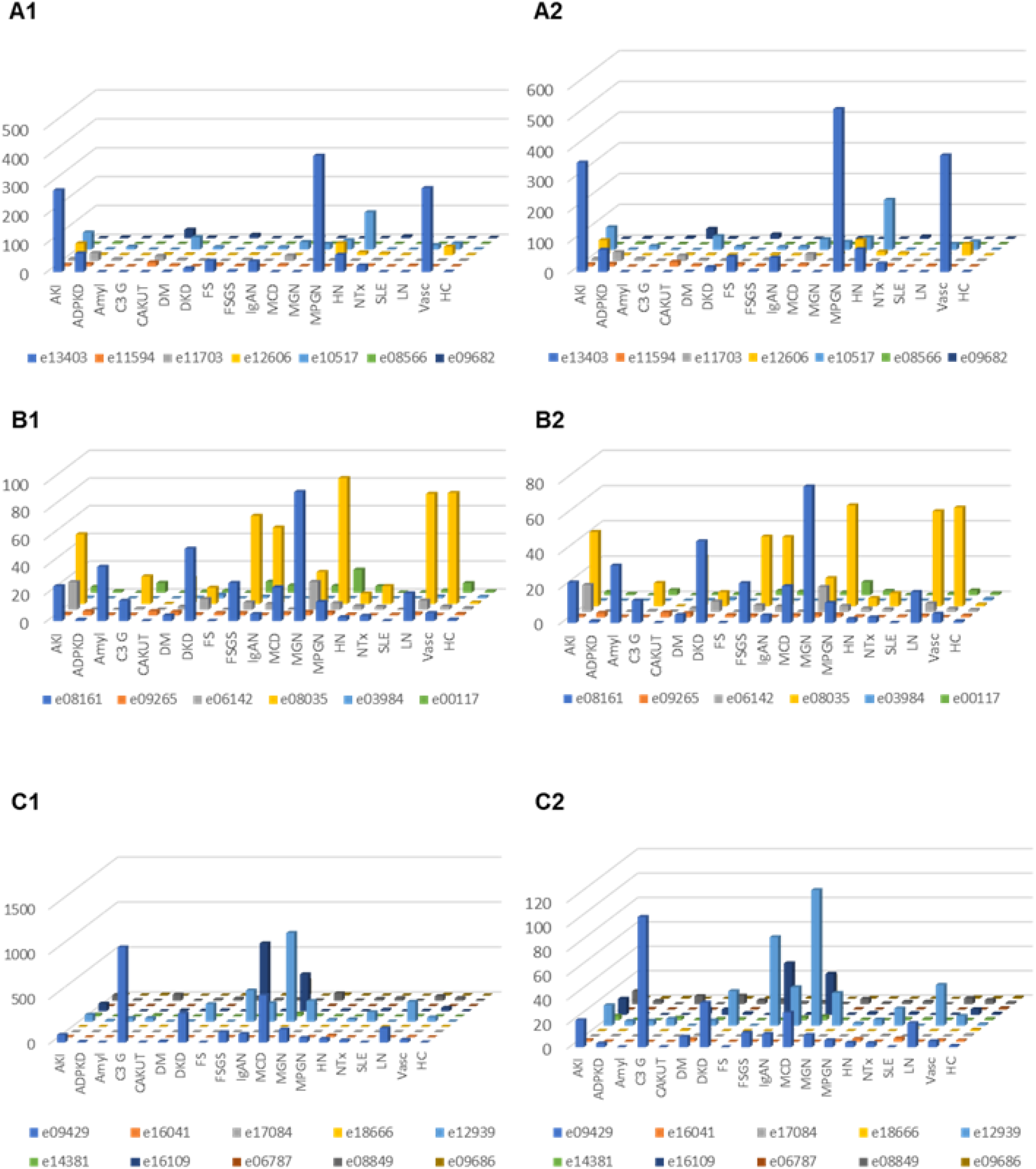
Relative abundances (normalized to the medium abundance in healthy controls) of complement fragments from the four different members of the complement family, unadjusted or adjusted for proteinuria. A1) Complement factor B derived peptides, unadjusted. A2) Complement factor B derived urine peptides, adjusted for proteinuria. B1) Peptides from complement factor 4A and 4B, unadjusted, B2) Complement factor 4A and 4B, adjusted for proteinuria. C1) Peptides from complement factor 3, unadjusted, C2) Peptides from complement factor 3, adjusted

## Discussion

This is the first study to describe clusters of complement fragment excretion in urine in a large cohort of patients with different kidney disease etiologies and/or clinical scenarios. Complement peptide excretions may serve as a surrogate of complement system activation status, possibly mirroring some of the processes at the cellular or circulation level.

There is very little data available on the urinary excretion of complement protein fragments in healthy individuals. The vast majority of complement factors have a molecular mass far above the threshold for glomerular filtration in the range of 15-50 kDa [16]. The larger complement proteins are c C1q (410kDa), C4 (210kDa), C4b (200kDa), C3 (180kDa) as well as C5 (180kDa). These complement factors are composed of polypeptide chains (usually of sizes between 22.5 and 40 kDa) [17]. Complement is activated by proteolysis, which leads to the generation of cleavage products, some of which pass through the glomerular basement membrane. However, this issue has not been systematically investigated yet. Low levels of complement C3 were detected in urine of healthy volunteers [18]. In this study the authors claimed that 90% of filtered complement C3 are reabsorbed at the proximal tubule [18]. Evidence exists on significant urinary complement C3 excretion in MPGN, MGN, IgAN and LN, similar to the findings in our investigation. There was a significant correlation between urinary excretion of complement C3 with intraglomerular immune complex deposition, especially along the glomerular capillary wall and less so when the deposits were solely at the mesangium [18].

In our study the extent of proteinuria was significantly and directly correlated with urinary C3 derived peptide excretion, while this was not observed for CFB. This is to some extent to be expected, as the plasma level of complement C3 is about 1.0 – 1.5 g/L, while CFB is present in much lower amount (ca 35 mg/L). Thus, proteinuria will lead to substantial amount of complement C3 in urine, while a 50-fold lower amount of urinary CFB as a result of proteinuria is expected. In fact, the minor yet significant inverse correlation of CFB peptide abundance with proteinuria may be due to a small, yet detectable signal suppression caused by the unspecific proteinuria.

When comparing urinary complement levels with those in plasma in different diseases, no significant correlation between levels of complement activation products in urine and in serum was found [19]. The levels of urinary complement (iC3b, Bb and complement membrane attack complex) were reported highest in FSGS and DKD, very low in minimal change nephropathy and almost undetectable in healthy individuals.

For most urine complement C3 fragment levels we observed an inverse association with eGFR. This inverse association was generally not observed for complement C4, while CFB fragments generally showed a positive correlation with eGFR (**Table 1**). Lower eGFR is associated with increased proteinuria in the dataset investigated (the association is shown in **supplementary Figure 3**). It seems plausible that the observed association of specific C3 complement fragments with eGFR is in fact the result of proteinuria. The data presented here support this assumption. However, no data are available to prove or reject this hypothesis with certainty. All of the CFB peptides originate from the same region and represent Bb fragments, being a part of the C3 convertase and constituting a stable complex with C3b on target surfaces. Smaller and more intensively processed Ba fragments were not detectable, presumably due to complete digestion during the complement activation.

When investigating the excretion of the most abundant peptide of C3 (LQGTPVAQMTEDAVDAERLKHL) in different diseases after normalization of individual excretion values to the median abundance of healthy controls (**Figure 2**), the highest relative levels were documented in MCD, FSGS, IgAN and LN, entities with known involvement of complement activation in the pathophysiology of the diseases. As depicted in **Figures 2, 3** and **4**, there is a remarkable high excretion of complement fragments in MGN. This intense complement excretion cluster in MGN may seem surprising and for a long time the disease has not been associated with increased complement activity. Nevertheless, evidence by now suggests that the complement system plays an important role in MGN [20], even though the complement pathways in MGN remain unexplored to some extent. Proteome analyses of kidney tissue samples from patients with membranous nephropathy also showed significant complement activation and evidence of complement peptides from C3 and C4-related pathways [21]. Urinary excretion of complement activation products (sC5b-9, C3d) has been shown to correlate with disease activity [22-24].

Complement peptides detected in urine do not cover the entire complement molecules, as would be expected if generation and secretion into urine was by chance and random. Instead, specific regions of the respective complement proteins are covered only, most pronounced for complement factor B, where all peptides detected are from the same region, AA positions 234-257. Upon closer examination of the regions represented from all three complement proteins, it appears that these are in each case regions that do not appear to have an active role in the complement activation, but rather are removed upon activation. As such, it is an attractive hypothesis that these peptides may in fact display previous complement activation, and could be exploited as biomarkers for such activation.

We found increased levels of most urinary complement peptides in LN in comparison to SLE, also after adjustment for proteinuria (**Figure 3** and **4**). Analysis of urinary complement excretion may be of interest in LN patients since urinary C3d was superior to plasma C3, C4d, Bb, C5b-9 and anti-double-stranded DNA antibody in distinguishing patients with LN from those without acute LN [25].

In IgAN there has been increasing appreciation of the complement system as mediator and contributor to the renal inflammation and tissue damage, but the mechanism of activation of the complement system and the contributing role of complement are not well defined and in part unclear [26;27]. In recent years the contributing role of the alternate and lectin pathway of complement activation have been introduced to the pathogenesis of IgAN. In our analysis we detected high levels of C3 (e12939) and CFB (e13403) fragments in urine (**Figure 2**), factors that can be attributed to the alternate pathway of complement activation. In the PerSTIgAN cohort [15], urinay C3 (e12939) excretion was significantly higher in patients exhibiting higher GFR loss (progressors) vs. those with lower GFR loss during follow-up (non-progressors).

In vasculitis urinary excretion of complement fragments was increased compared to healthy controls. The majority of patients in this group had renal involvement of anti-neutrophil cytoplasmic antibody (ANCA)-associated vasculitis. Despite the microscopic “pauci-immune” nature of the glomerulonephritis in immunofluorescence evaluation, the available evidence indicates that activation of the complement system through the alternative pathway is necessary for the development of ANCA-associated vasculitis and is also associated with prognosis of the disease [28].

In patients with DKD, complement fragment excretion was higher than in healthy controls, but also higher than in diabetic patients with preserved renal function (**Figure 2, 3 and 4**). Especially excretion of C3 complement fragments was higher in patients with DKD than in diabetic patients without kidney involvement. The important role of complement activation in DKD and its importance for renal pathology and the progression of kidney damage is becoming increasingly evident [29;30].

Transcriptome and immunohistochemical analyses of human and mouse kidney biopsies of the diseases investigated in the current study revealed increased glomerular and/or tubular deposition of C3 and CFB compared to healthy controls [31-34]. C4A mRNA was reported increased in glomeruli of DKD patients compared to healthy controls [31]. These data reinforce the hypothesis that complement activation contributes to the pathogenesis of a variety of kidney diseases and potentially induces tissue damage through inflammation.

A growing body of data from mouse experimental studies demonstrate the importance of complement cascade in kidney diseases. Glomerular C3 deposits have been detected increased in T1DM non-obese diabetic mouse [35] and in OVE26 diabetic mouse [36]. C3 and C3 cleaved products were detected in the glomeruli of adriamycin-treated mice mimicking human FSGS [37]. C4 was associated with LN mouse models [38] As such, the human data from this study could also be used as a basis to investigate complement and its fragments in animal models in more detail.

Despite the positive correlation of complement components in urine of several complement mediated kidney disorders, this study has several limitations. First, we could not control for confounder that might influence complement activation and complement peptide excretion (e.g. infections, different type of diseases in the vasculitis group, different types of LN, heterogenous variants of FSGS, therapeutic interventions) in the different subcategories of diseases. Second, we did not have values for proteinuria and eGFR available for all patients in the study. Nevertheless, the still large number of datasets with associated proteinuria and/or eGFR certainly represents a major strength of the report. Third, the sole urinary output of a complement fragment does not automatically associate with a contributing role in disease processes. As an example, the C4d deposition, an important diagnostic tool in antibody mediated rejections of renal allografts, does not correlate with urinary C4d excretion, consequently C4d urine levels are of no value as biomarkers in AMR [39].

In conclusion, we provide evidence for the activation-excretion of complement components in urine samples of several human kidney diseases. Multiple complement-derived peptides were significantly associated with kidney function, and with specific kidney disease aetiologies. Proteomic screening of these complement factors in urine could provide a basis for assessing kidney disease. Complement fragments derived from C3, C4 and CFB show distinct patterns in different diseases. They might provide useful information on status of disease, to predict and/or assess response to therapy, and might serve as a diagnostic tool, e.g. discrimination between active and non-active disease. Based on the data available, it appears plausible that urinary complement fragments are associated with different CKD etiology/ condition and could potentially guide therapeutic decisions in a personalized medicine approach.

## Data Availability

Data can be requested at corresponding author

## Acknowledgement

The work of PFZ and TW is supported by the Collaborative Research Center (CRC) SFB 1192 by the Deutsche Forschungsgemeinschaft (DFG), project B6, Kidneeds (PFZ), and the Bundesministerium für Bildung und Forschung (BMBF), DEFEAT PANDEMIcs (TW)

## Statement of Ethics

The work presented in that manuscript complies with guidelines for human studies. Existing anonymized datasets from ethical approved published studies were employed.

## Data sharing

Anonymised data will be made available upon request directed to the corresponding author. Proposals will be reviewed and approved by the investigators and collaborators based on scientific merit. After approval of a proposal, data will be shared through a secure online platform after signing a data access and confidentiality agreement.

## Supplementary Material

**Supplementary Figure 1:** Tandem mass spectra and assignment of sequences of the 23 complement derived urine peptides detected.

**Supplementary Figure 2:** Association of the combined abundance of complement peptides (from Complement factor B, 4A, 4B and 3) with proteinuria, A) prior and B) after adjustment for proteinuria.

**Supplementary Figure 3:** Association of proteinuria with eGFR in the study population. As expected, a highly significant correlation is detectable.

**Supplementary Table 1** Relative abundance of all 23 collagen derived peptides, listed per disease group, normalized to the normal controls.

