## supplement for "Molecular mapping of urinary complement peptides in kidney diseases"

**e-Supplement** to

**Differential excretion of urinary complement fragments in kidney diseases: a potential link to complement activation?**

### Supplementary Figure 1 (part 1-23)

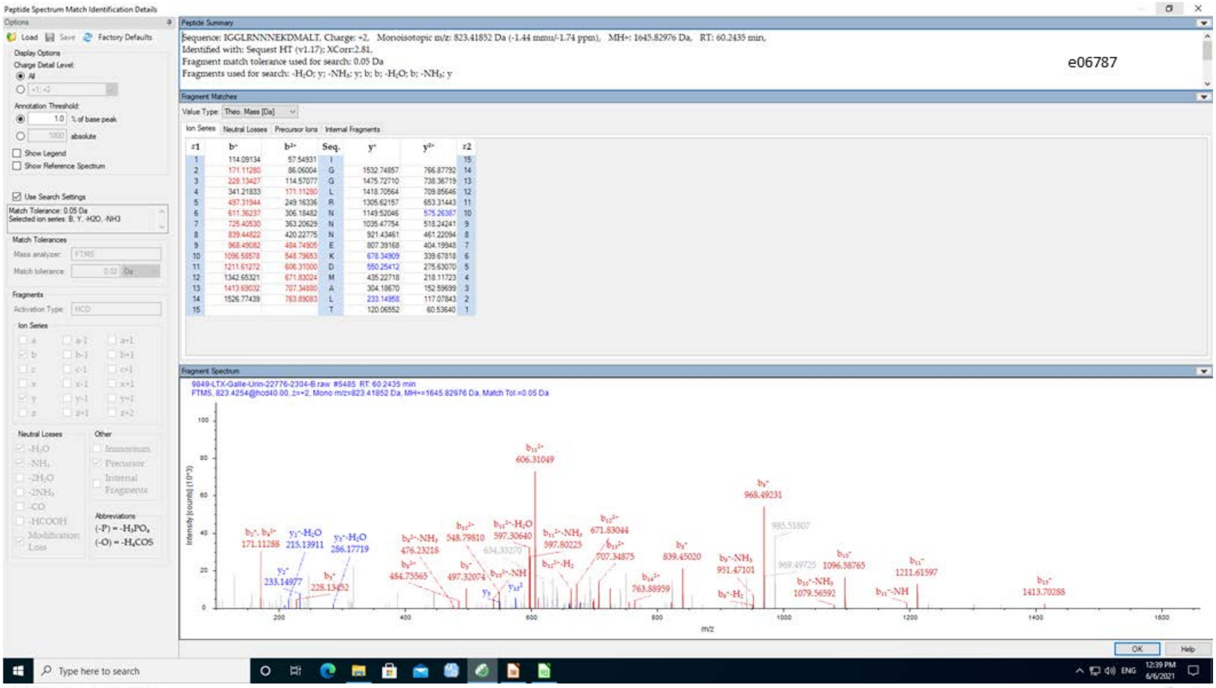


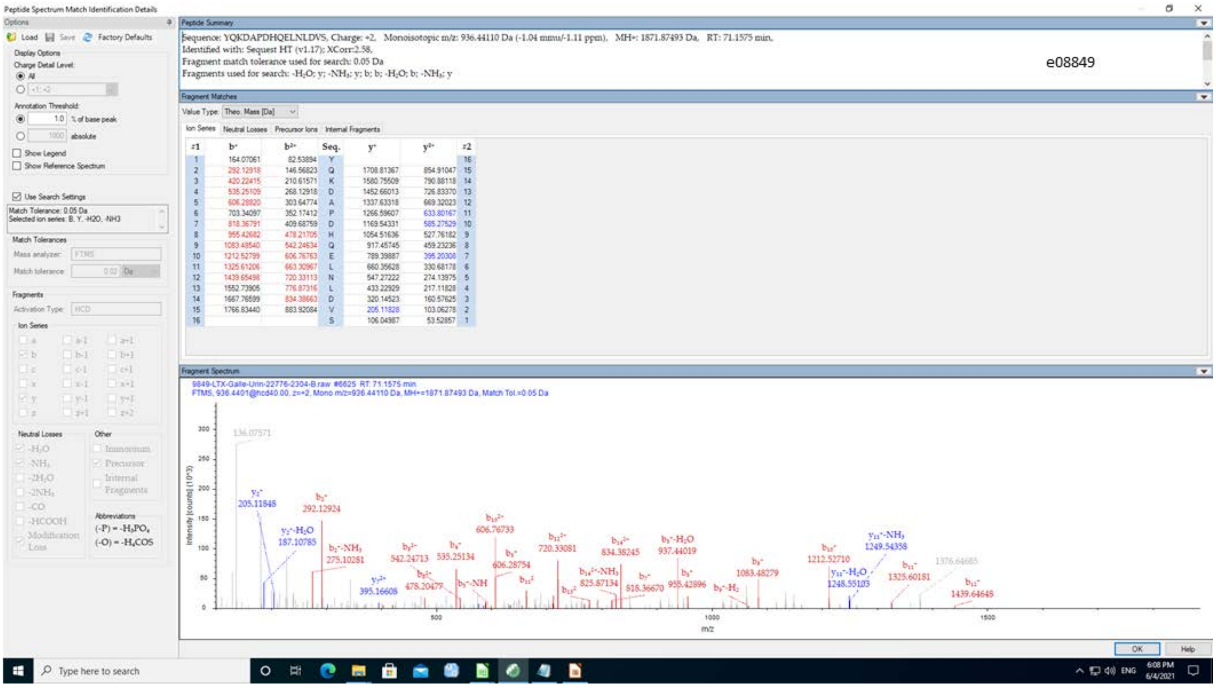

e09686

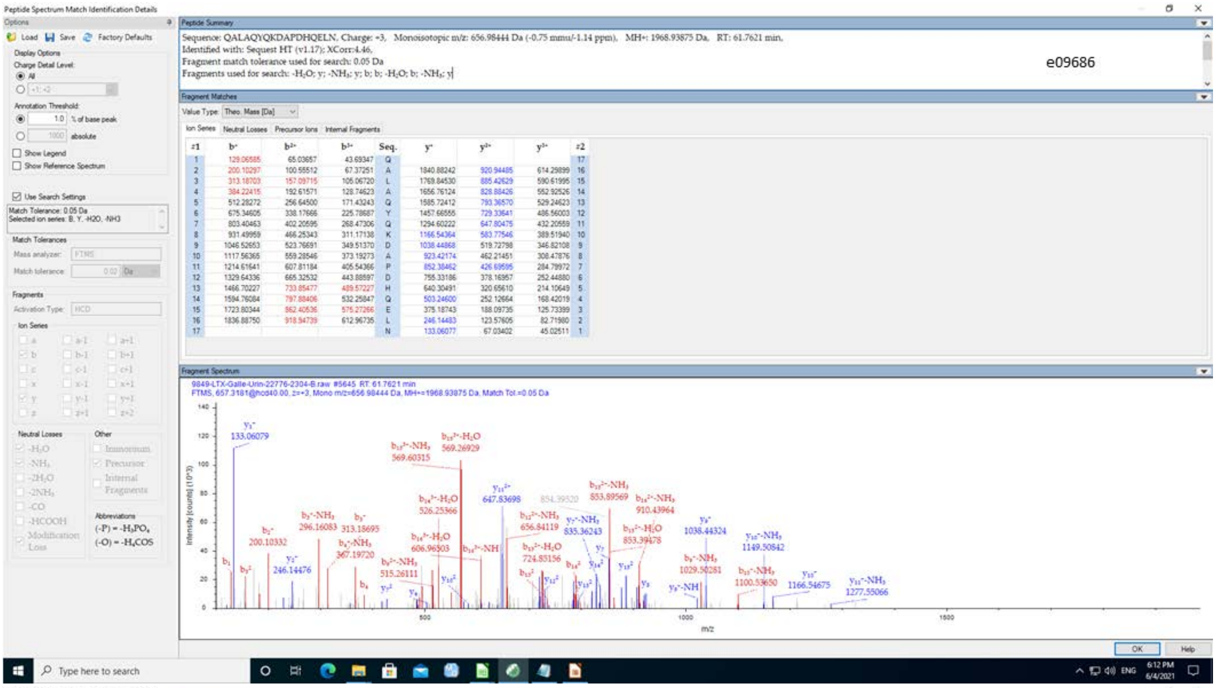


| #1 | b+ | b2+ | b3+ | Seq. | y+ | y2+ | y3+ | #2 |
|---|---|---|---|---|---|---|---|---|
| 1 | 129.06585 | 65.03657 | 43.69347 | Q | | | | 17 |
| 2 | 200.10297 | 100.55512 | 67.37251 | A | 1840.88242 | 920.94485 | 614.29899 | 16 |
| 3 | 313.18703 | 157.09715 | 105.06720 | L | 1769.84530 | 885.42629 | 590.61995 | 15 |
| 4 | 384.22415 | 192.61571 | 128.74623 | A | 1656.76124 | 828.88426 | 552.92526 | 14 |
| 5 | 512.28272 | 256.64500 | 171.43243 | Q | 1585.72412 | 793.36570 | 529.24623 | 13 |
| 6 | 675.34605 | 338.17666 | 225.78687 | Y | 1457.66555 | 729.33641 | 486.56003 | 12 |
| 7 | 803.40463 | 402.20595 | 268.47306 | Q | 1294.60222 | 647.80475 | 432.20559 | 11 |
| 8 | 931.49959 | 466.25343 | 311.17138 | K | 1166.54364 | 583.77546 | 389.51940 | 10 |
| 9 | 1046.52653 | 523.76691 | 349.51370 | D | 1038.44868 | 519.72798 | 346.82108 | 9 |
| 10 | 1117.56365 | 559.28546 | 373.19273 | A | 923.42174 | 462.21451 | 308.47876 | 8 |
| 11 | 1214.61641 | 607.81184 | 405.54366 | P | 852.38462 | 426.69595 | 284.79972 | 7 |
| 12 | 1329.64336 | 665.32532 | 443.88597 | D | 755.33186 | 378.16957 | 252.44880 | 6 |
| 13 | 1466.70227 | 733.85477 | 489.57227 | H | 640.30491 | 320.65610 | 214.10649 | 5 |
| 14 | 1594.76084 | 797.88406 | 532.25847 | Q | 503.24600 | 252.12664 | 168.42019 | 4 |
| 15 | 1723.80344 | 862.40536 | 575.27266 | E | 375.18743 | 188.09735 | 125.73399 | 3 |
| 16 | 1836.88750 | 918.94739 | 612.96735 | L | 246.14483 | 123.57605 | 82.71980 | 2 |
| 17 | | | | N | 133.06077 | 67.03402 | 45.02511 | 1 |

e14381

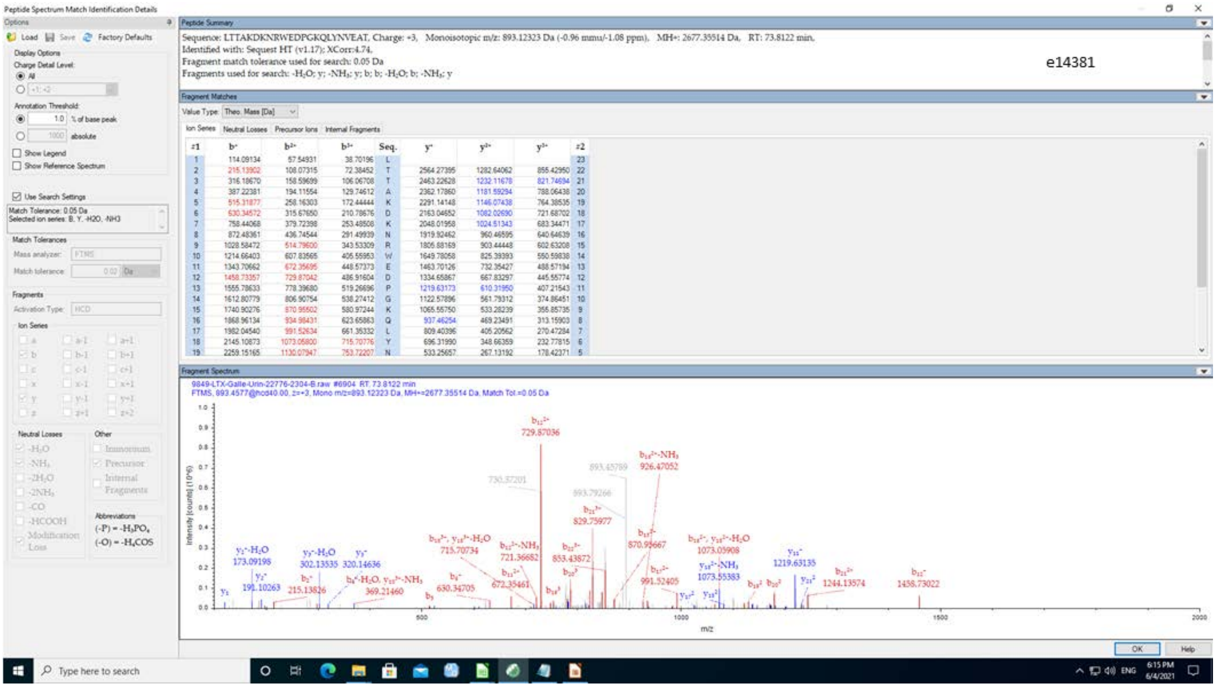


| #1 | b+ | b2+ | b3+ | Seq. | y+ | y2+ | y3+ | #2 |
|---|---|---|---|---|---|---|---|---|
| 1 | 114.09134 | 57.54931 | 38.70196 | L | | | | 23 |
| 2 | 215.13902 | 108.07315 | 72.38452 | T | 2564.27395 | 1282.64062 | 855.42950 | 22 |
| 3 | 316.18670 | 158.59699 | 106.06708 | T | 2463.22628 | 1232.11678 | 821.74694 | 21 |
| 4 | 387.22381 | 194.11554 | 129.74612 | A | 2362.17860 | 1181.59294 | 788.06438 | 20 |
| 5 | 515.31877 | 258.16303 | 172.44444 | K | 2291.14148 | 1146.07438 | 764.38535 | 19 |
| 6 | 630.34572 | 315.67650 | 210.78676 | D | 2163.04652 | 1082.02690 | 721.68702 | 18 |
| 7 | 758.44068 | 379.72398 | 253.48508 | K | 2048.01958 | 1024.51343 | 683.34471 | 17 |
| 8 | 872.48361 | 436.74544 | 291.49939 | N | 1919.92462 | 960.46595 | 640.64639 | 16 |
| 9 | 1028.58472 | 514.79600 | 343.53309 | R | 1805.88169 | 903.44448 | 602.63208 | 15 |
| 10 | 1214.66403 | 607.83565 | 405.55953 | W | 1649.78058 | 825.39393 | 550.59838 | 14 |
| 11 | 1343.70662 | 672.35695 | 448.57373 | E | 1463.70126 | 732.35427 | 488.57194 | 13 |
| 12 | 1458.73357 | 729.87042 | 486.91604 | D | 1334.65867 | 667.83297 | 445.55774 | 12 |
| 13 | 1555.78633 | 778.39680 | 519.26696 | P | 1219.63173 | 610.31950 | 407.21543 | 11 |
| 14 | 1612.80779 | 806.90754 | 538.27412 | G | 1122.57896 | 561.79312 | 374.86451 | 10 |
| 15 | 1740.90276 | 870.95502 | 580.97244 | K | 1065.55750 | 533.28239 | 355.85735 | 9 |
| 16 | 1868.96134 | 934.98431 | 623.65863 | Q | 937.46254 | 469.23491 | 313.15903 | 8 |
| 17 | 1982.04540 | 991.52634 | 661.35332 | L | 809.40396 | 405.20562 | 270.47284 | 7 |
| 18 | 2145.10873 | 1073.05800 | 715.70776 | Y | 696.31990 | 348.66359 | 232.77815 | 6 |
| 19 | 2259.15165 | 1130.07947 | 753.72207 | N | 533.25657 | 267.13192 | 178.42371 | 5 |

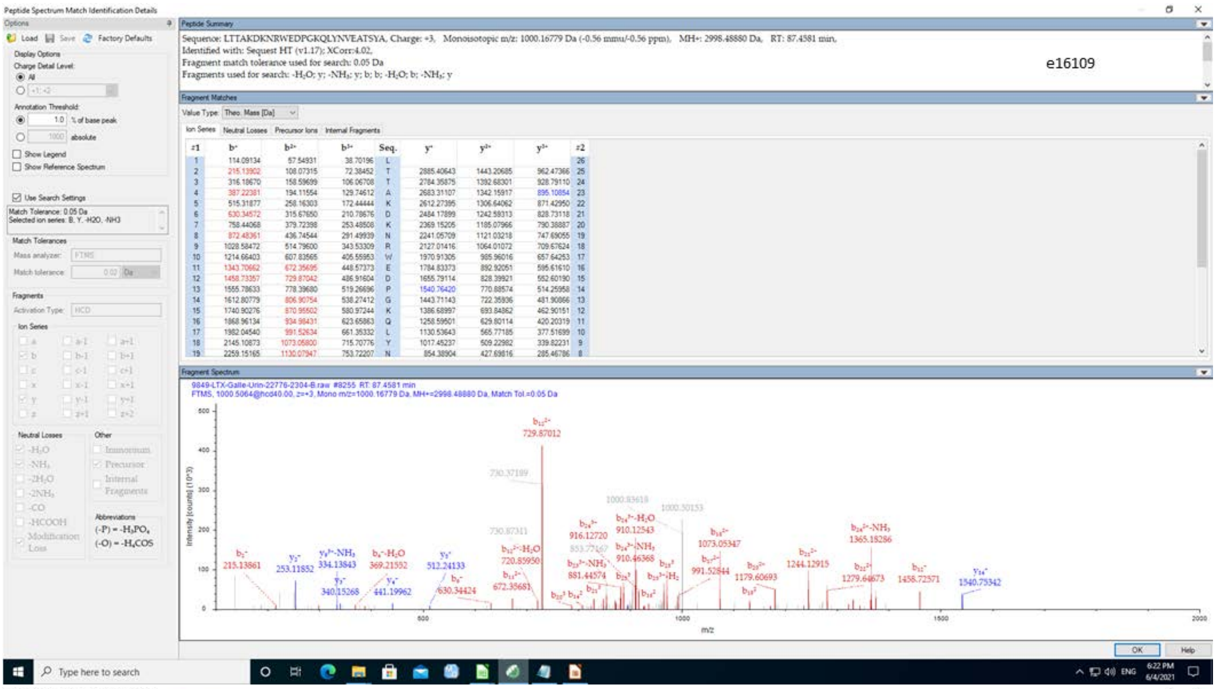
Peptide Spectrum Match Identification Details
e16109
Sequence: LTTAKDKNRWEDPGKQLYNVEATSYA, Charge: +3, Monoisotopic m/z: 1000.16779 Da (-0.56 mmu/-0.56 ppm), MH+: 2998.48880 Da, RT: 87.4581 min,
Identified with: Sequest HT (v1.17); XCorr:4.02,
Fragment match tolerance used for search: 0.05 Da
Fragments used for search: -H2O; y; -NH3; y; b; b; -H2O; b; -NH3; y
Fragment Matches
Fragment Spectrum
m/z

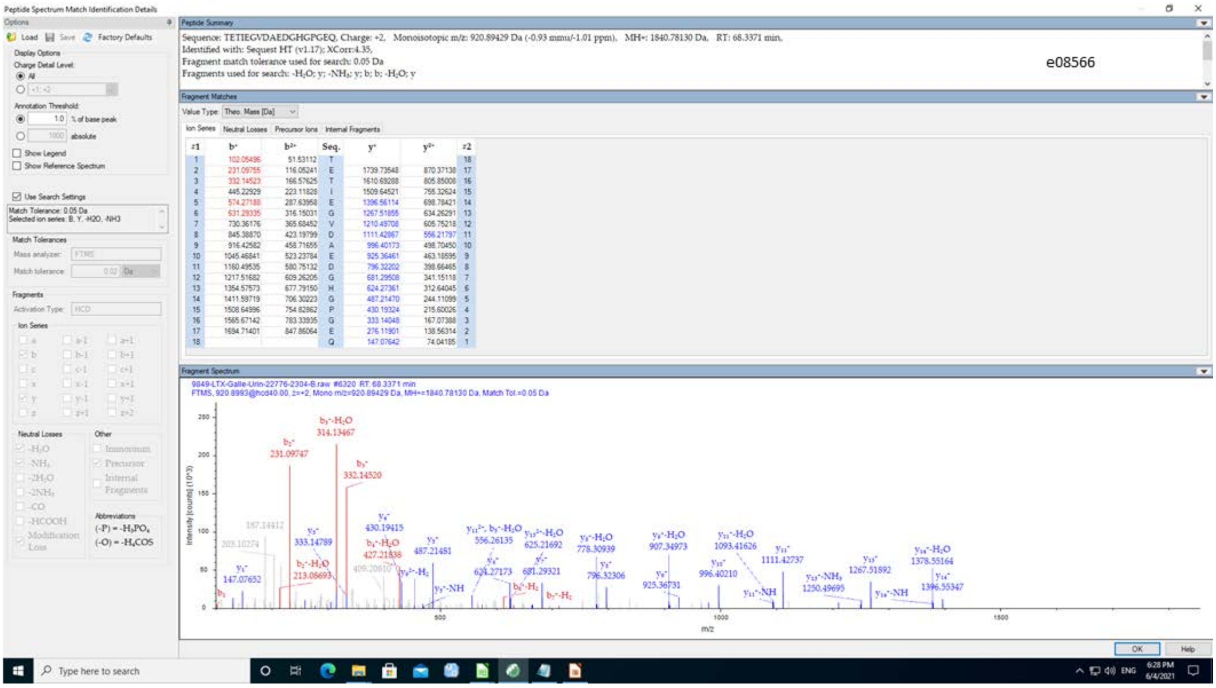
Peptide Spectrum Match Identification Details
e08566
Sequence: TETIEGVDAEDGHGPGEQ, Charge: +2, Monoisotopic m/z: 920.89429 Da (-0.93 mmu/-1.01 ppm), MH+: 1840.78130 Da, RT: 68.3371 min,
Identified with: Sequest HT (v1.17); XCorr:4.35,
Fragment match tolerance used for search: 0.05 Da
Fragments used for search: -H2O; y; -NH3; y; b; b; -H2O; y
Fragment Matches
Fragment Spectrum
m/z

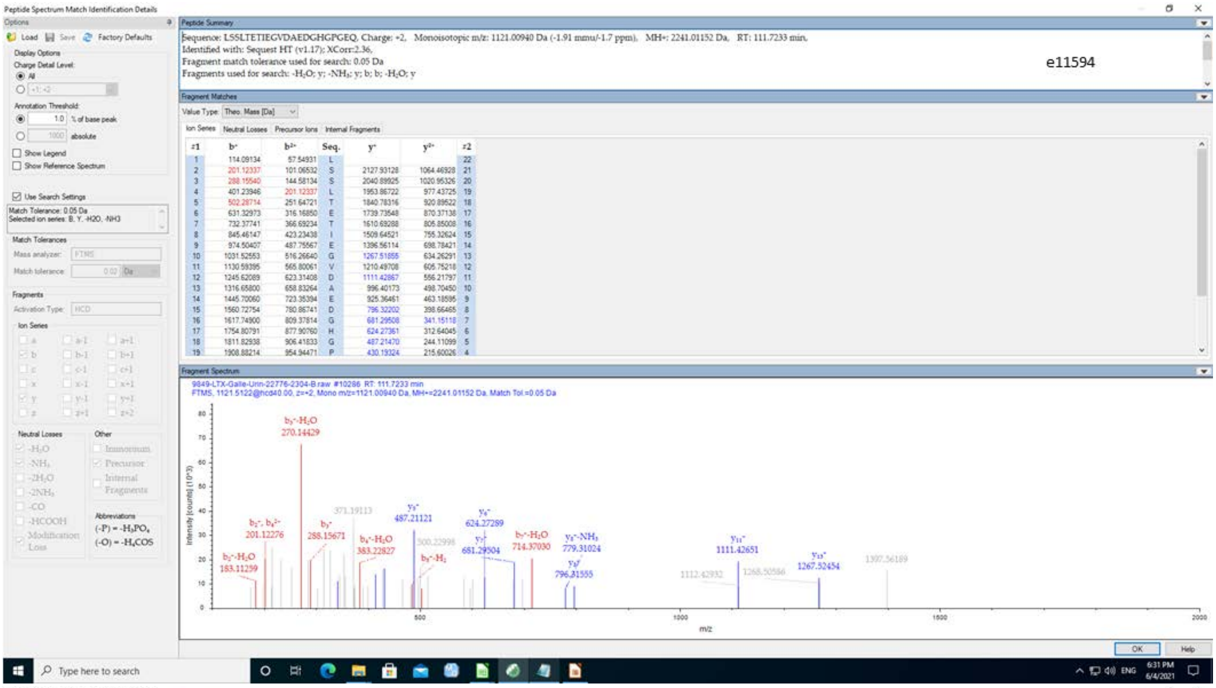

Peptide Spectrum Match Identification Details
Sequence: LSSLTETIEGVDAEDGHGPGEQ, Charge: +2, Monoisotopic m/z: 1121.00940 Da (-1.91 mmu/-1.7 ppm), MH+: 2241.01152 Da, RT: 111.7233 min,
Identified with: Sequest HT (v1.17); XCorr:2.36,
Fragment match tolerance used for search: 0.05 Da
Fragments used for search: -H2O; y; -NH3; y; b; b; -H2O; y
e11594


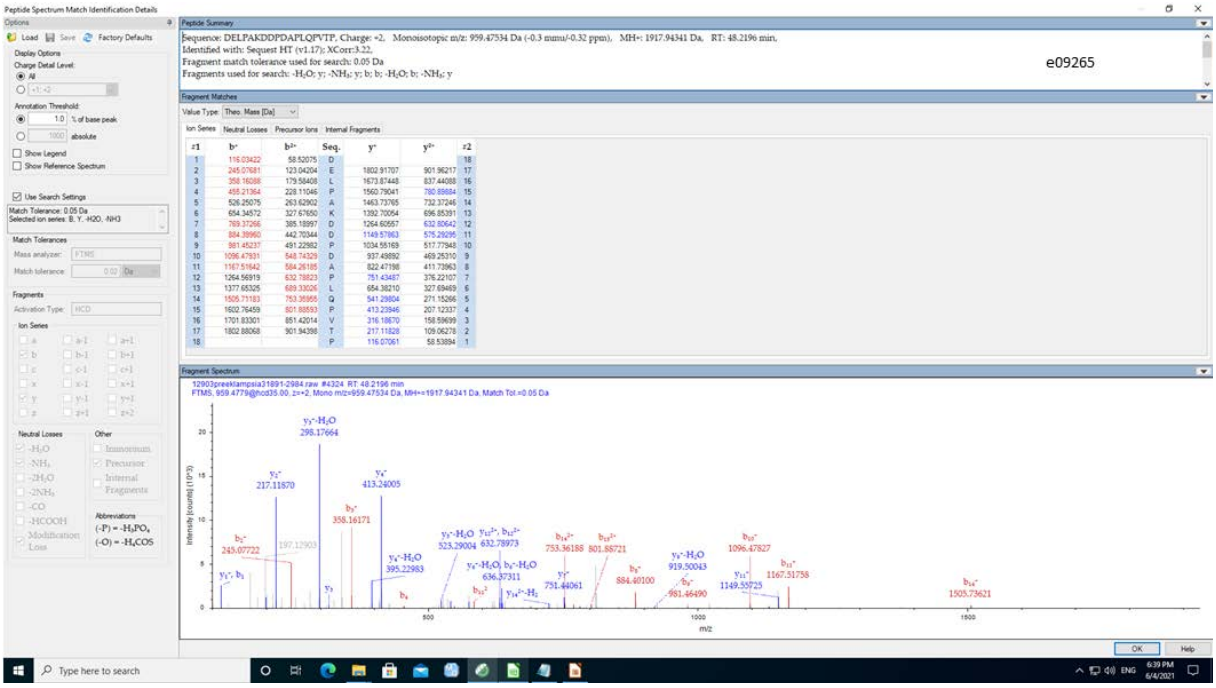

Peptide Spectrum Match Identification Details
Sequence: DELPAKDDPDAPLQPVTP, Charge: +2, Monoisotopic m/z: 959.47534 Da (-0.3 mmu/-0.32 ppm), MH+: 1917.94341 Da, RT: 48.2196 min,
Identified with: Sequest HT (v1.17); XCorr:3.22,
Fragment match tolerance used for search: 0.05 Da
Fragments used for search: -H2O; y; -NH3; y; b; b; -H2O; b; -NH3; y
e09265

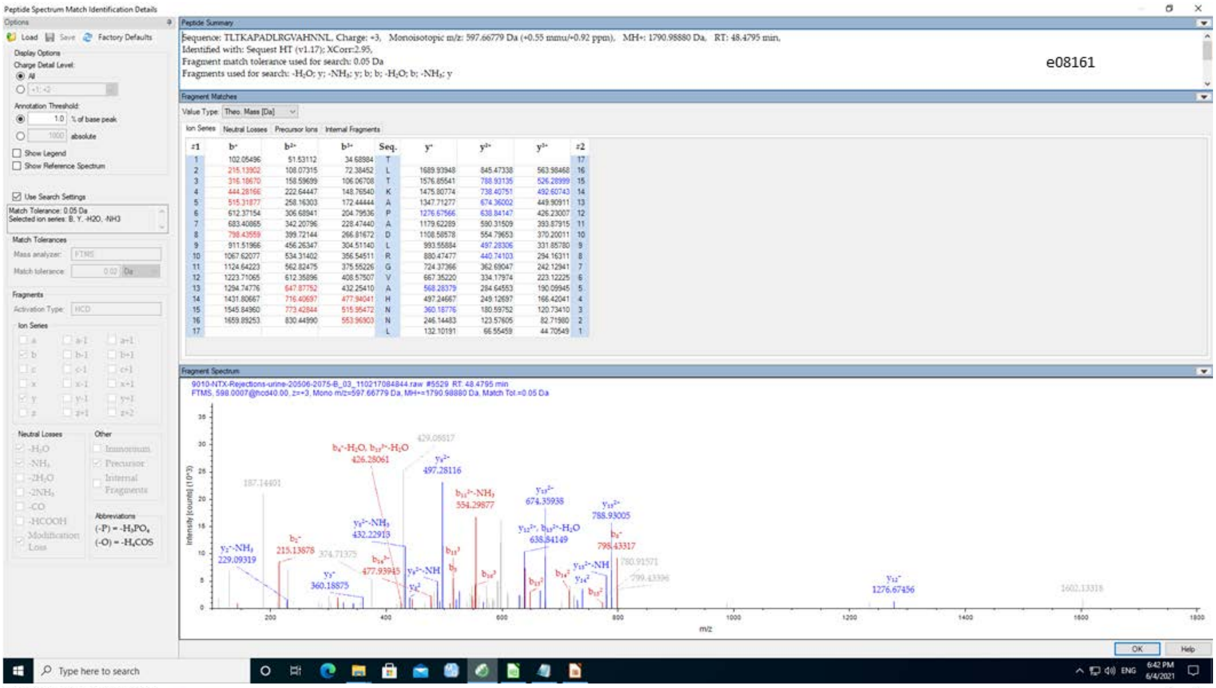

Peptide Spectrum Match Identification Details
Sequence: TLTKAPADLRGVAHNNL, Charge: +3, Monoisotopic m/z: 597.66779 Da (+0.55 mmu/+0.92 ppm), MH+: 1790.98880 Da, RT: 48.4795 min,
Identified with: Sequest HT (v1.17); XCorr:2.95,
Fragment match tolerance used for search: 0.05 Da
Fragments used for search: -H₂O; y; -NH₃; y; b; b; -H₂O; b; -NH₃; y
e08161


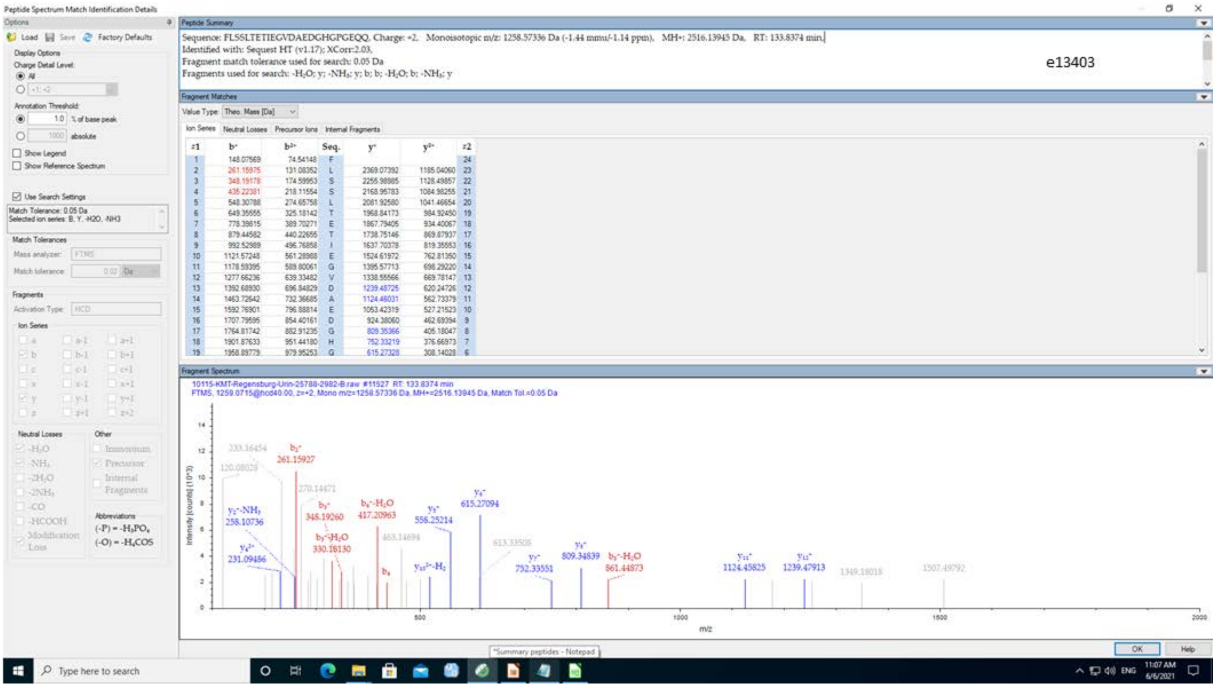

Peptide Spectrum Match Identification Details
Sequence: FLSSLTETIEGVDAEDGHGPGEQQ, Charge: +2, Monoisotopic m/z: 1258.57336 Da (-1.44 mmu/-1.14 ppm), MH+: 2516.13945 Da, RT: 133.8374 min,
Identified with: Sequest HT (v1.17); XCorr:2.03,
Fragment match tolerance used for search: 0.05 Da
Fragments used for search: -H₂O; y; -NH₃; y; b; b; -H₂O; b; -NH₃; y
e13403

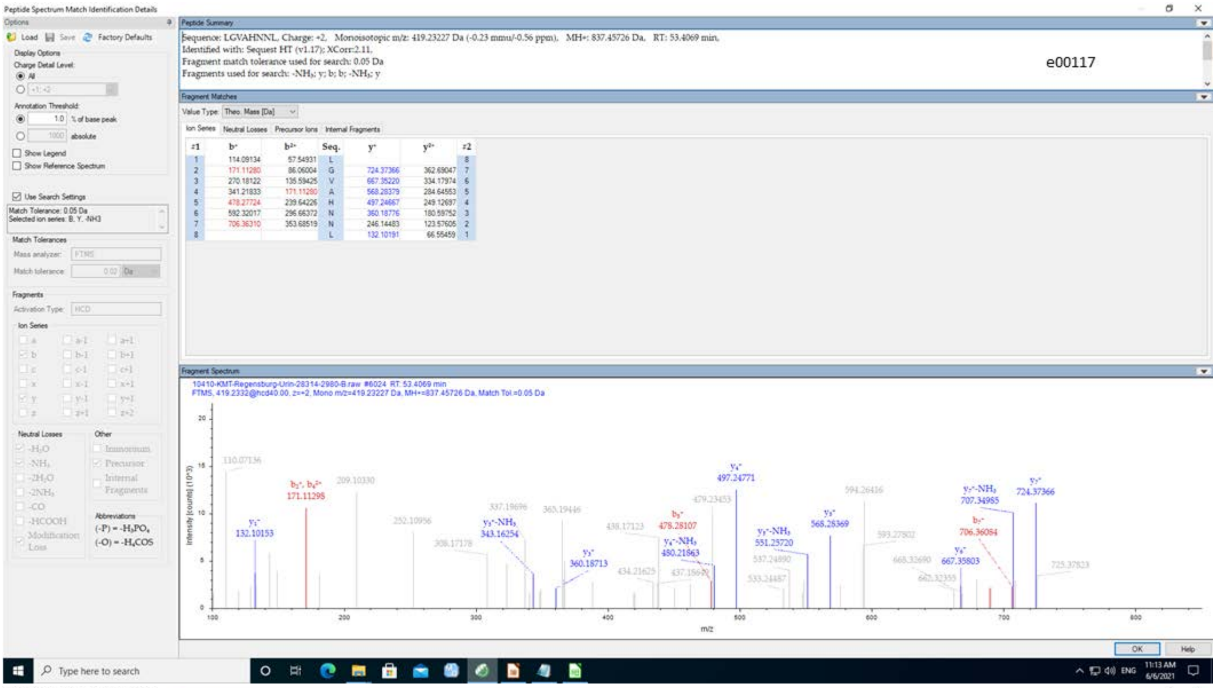
Peptide Spectrum Match Identification Details
Peptide Summary
Sequence: LGVAHNNL, Charge: +2, Monoisotopic m/z: 419.23227 Da (-0.23 mmu/-0.56 ppm), MH+: 837.45726 Da, RT: 53.4069 min,
Identified with: Sequest HT (v1.17); XCorr:2.11,
Fragment match tolerance used for search: 0.05 Da
Fragments used for search: -NH3; y; b; b; -NH3; y
e00117
Fragment Matches
Fragment Spectrum
m/z


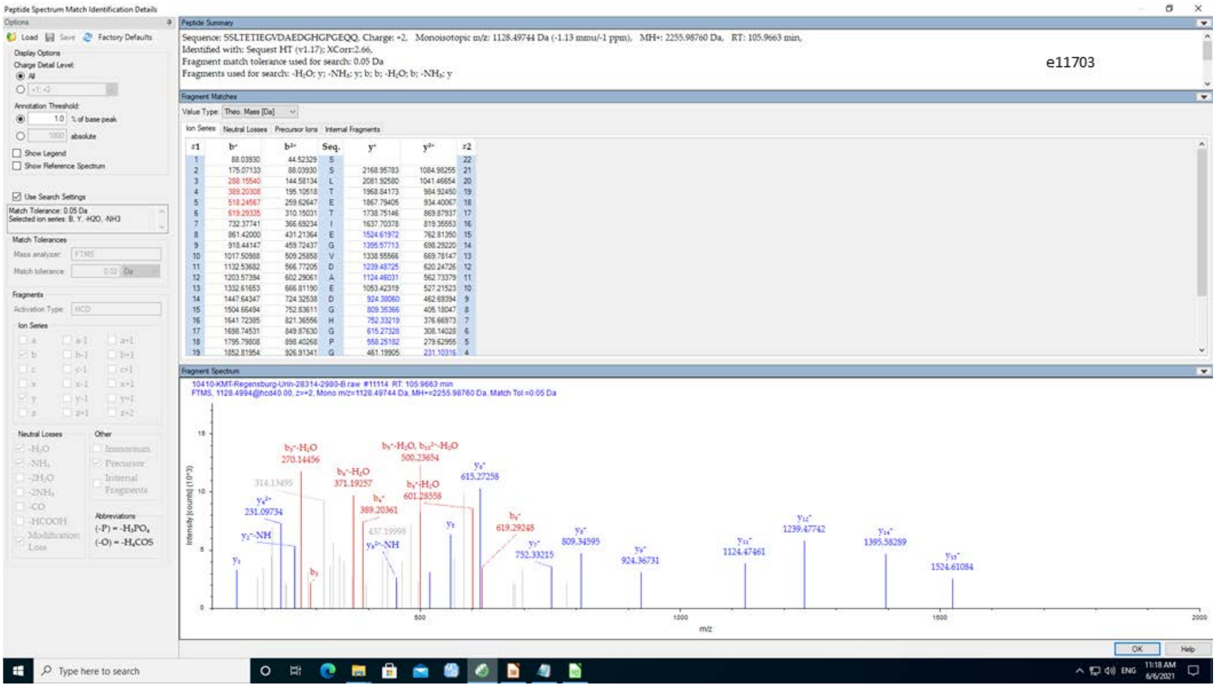
Peptide Spectrum Match Identification Details
Peptide Summary
Sequence: SSLTETIEGVDAEDGHGPGEQQ, Charge: +2, Monoisotopic m/z: 1128.49744 Da (-1.13 mmu/-1 ppm), MH+: 2255.98760 Da, RT: 105.9663 min,
Identified with: Sequest HT (v1.17); XCorr:2.66,
Fragment match tolerance used for search: 0.05 Da
Fragments used for search: -H2O; y; -NH3; y; b; b; -H2O; b; -NH3; y
e11703
Fragment Matches
Fragment Spectrum
m/z

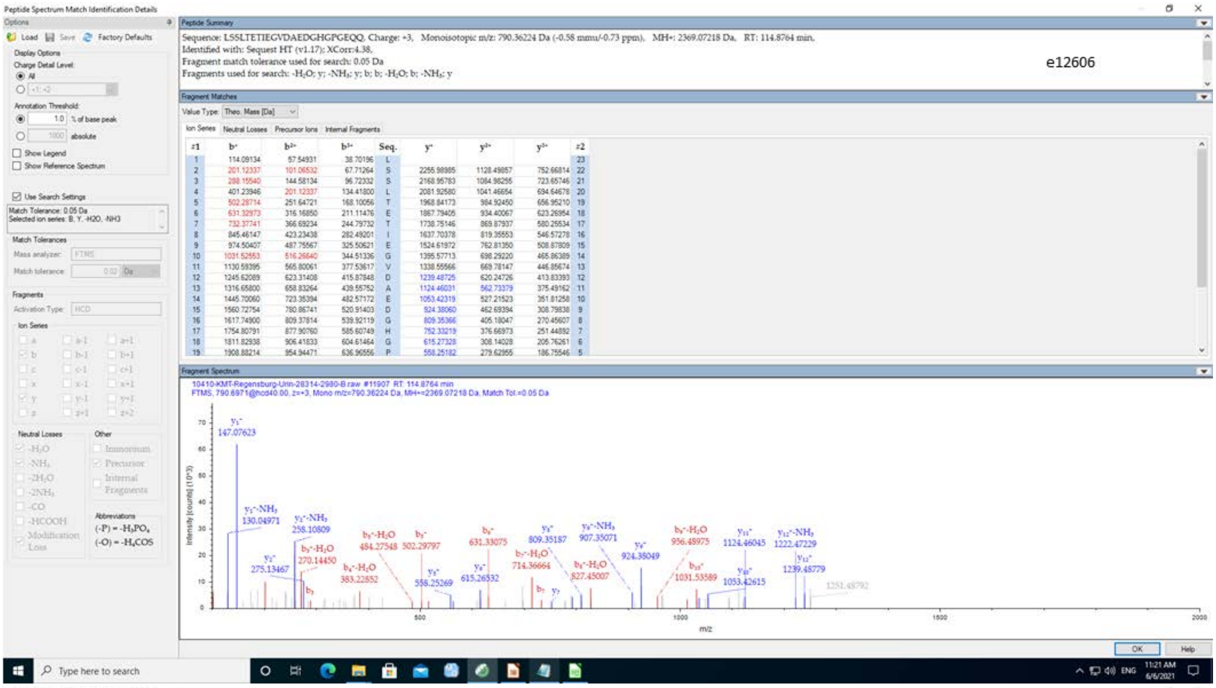
Peptide Spectrum Match Identification Details
Sequence: LSSLTETIEGVDAEDGHGPGEQQ, Charge: +3, Monoisotopic m/z: 790.36224 Da (-0.58 mmu/-0.73 ppm), MH+: 2369.07218 Da, RT: 114.8764 min,
Identified with: Sequest HT (v1.17); XCorr:4.38,
Fragment match tolerance used for search: 0.05 Da
Fragments used for search: -H2O; y; -NH3; y; b; b; -H2O; b; -NH3; y
e12606
10410-KMT-Regensburg-Urin-28314-2980-B.raw #11907 RT: 114.8764 min
FTMS, 790.6971@hcd40.00, z=+3, Mono m/z=790.36224 Da, MH+=2369.07218 Da, Match Tol.=0.05 Da

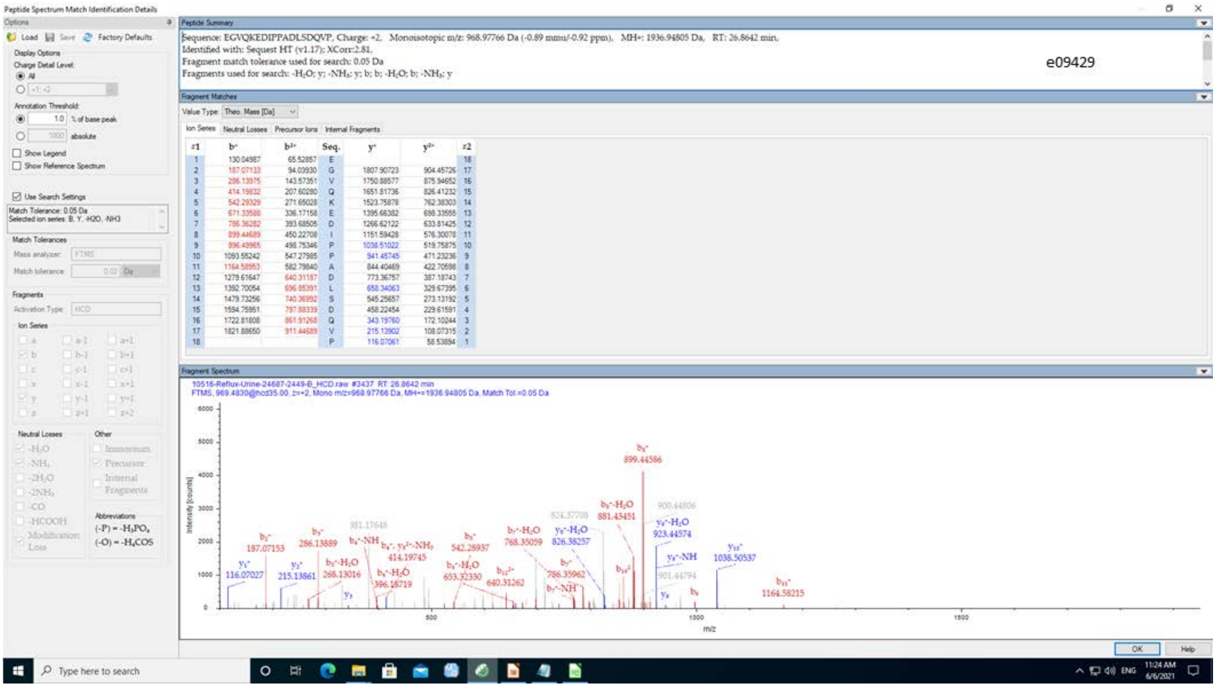
Peptide Spectrum Match Identification Details
Sequence: EGVQKEDIPPADLSDQVP, Charge: +2, Monoisotopic m/z: 968.97766 Da (-0.89 mmu/-0.92 ppm), MH+: 1936.94805 Da, RT: 26.8642 min,
Identified with: Sequest HT (v1.17); XCorr:2.81,
Fragment match tolerance used for search: 0.05 Da
Fragments used for search: -H2O; y; -NH3; y; b; b; -H2O; b; -NH3; y
e09429
10516-Reflux-Urine-24687-2449-B_HCD.raw #3437 RT: 26.8642 min
FTMS, 969.4830@hcd35.00, z=+2, Mono m/z=968.97766 Da, MH+=1936.94805 Da, Match Tol.=0.05 Da

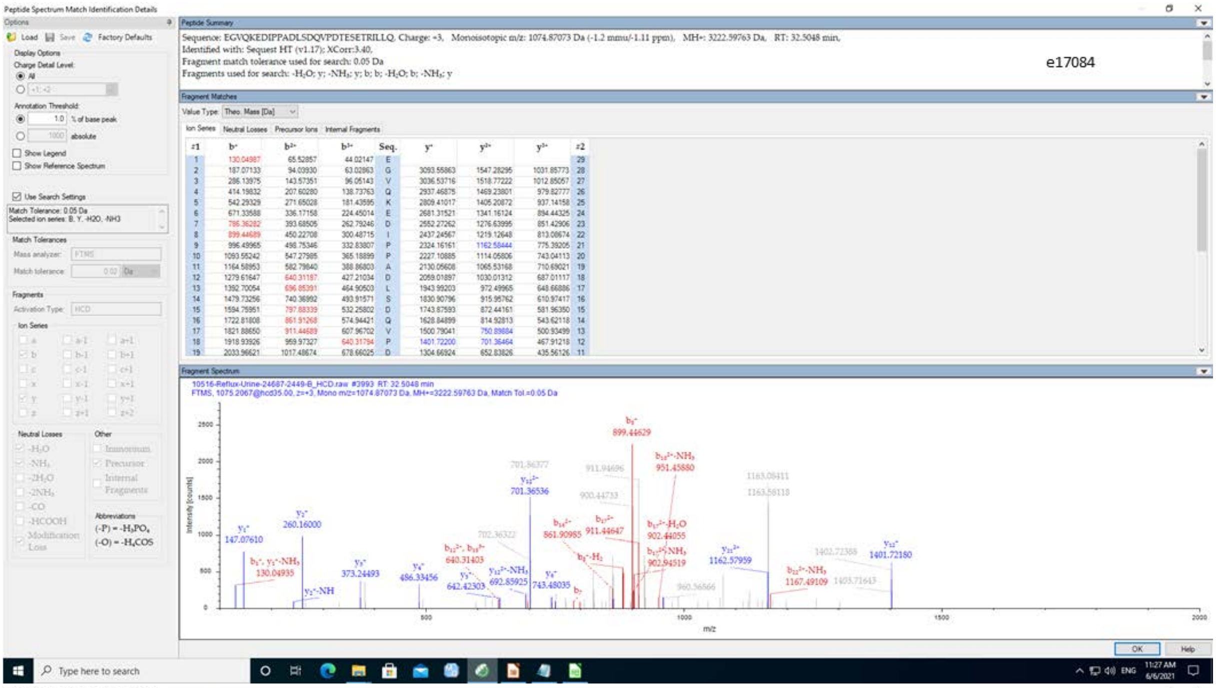
Peptide Spectrum Match Identification Details
Peptide Summary
Sequence: EGVQKEDIPPADLSDQVPDTESETRILLQ, Charge: +3, Monoisotopic m/z: 1074.87073 Da (-1.2 mmu/-1.11 ppm), MH+: 3222.59763 Da, RT: 32.5048 min,
Identified with: Sequest HT (v1.17); XCorr:3.40,
Fragment match tolerance used for search: 0.05 Da
Fragments used for search: -H2O; y; -NH3; y; b; b; -H2O; b; -NH3; y
e17084
Fragment Matches
Fragment Spectrum
m/z
Intensity [counts]

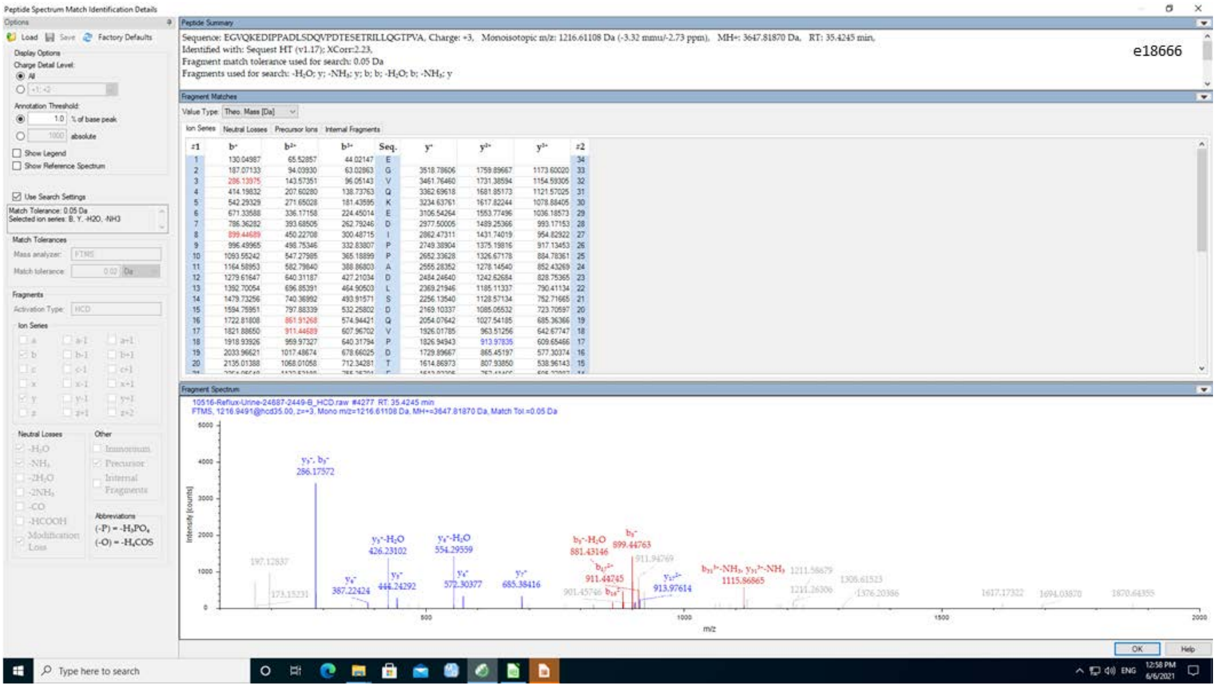
Peptide Spectrum Match Identification Details
Peptide Summary
Sequence: EGVQKEDIPPADLSDQVPDTESETRILLQGTPVA, Charge: +3, Monoisotopic m/z: 1216.61108 Da (-3.32 mmu/-2.73 ppm), MH+: 3647.81870 Da, RT: 35.4245 min,
Identified with: Sequest HT (v1.17); XCorr:2.23,
Fragment match tolerance used for search: 0.05 Da
Fragments used for search: -H2O; y; -NH3; y; b; b; -H2O; b; -NH3; y
e18666
Fragment Matches
Fragment Spectrum
m/z
Intensity [counts]

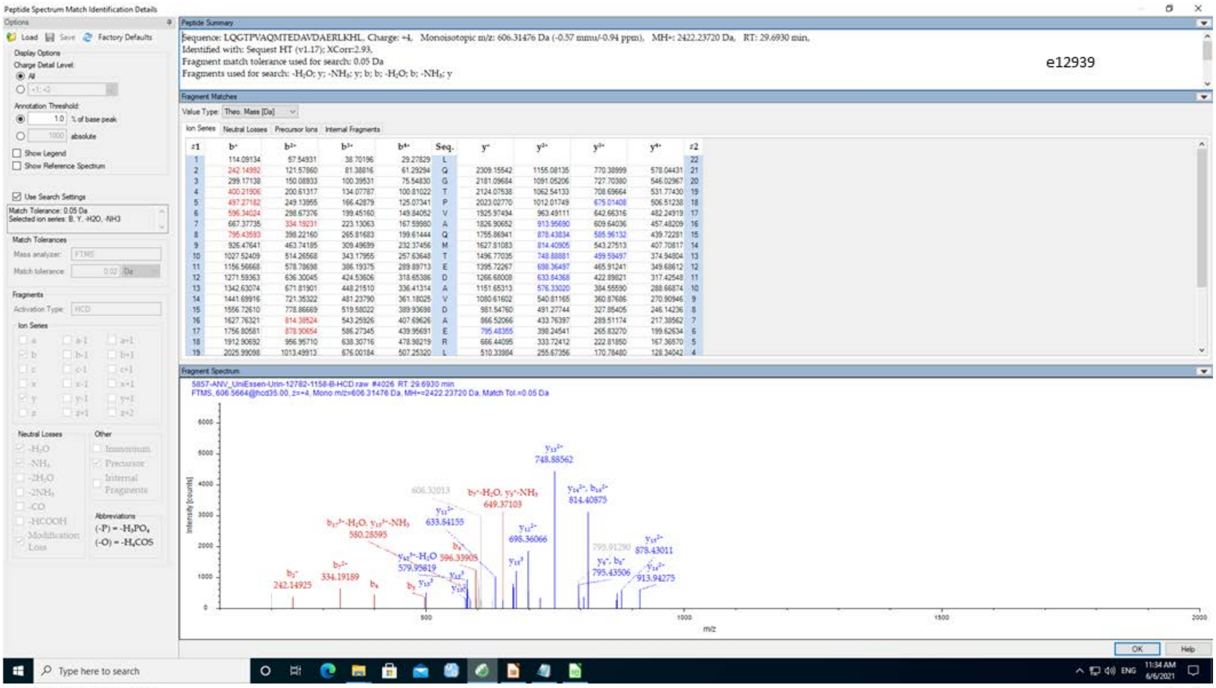
e12939
Sequence: LQGTPVAQMTEDAVDAERLKHL, Charge: +4, Monoisotopic m/z: 606.31476 Da (-0.57 mmu/-0.94 ppm), MH+: 2422.23720 Da, RT: 29.6930 min,
Identified with: Sequest HT (v1.17); XCorr:2.93,
Fragment match tolerance used for search: 0.05 Da
Fragments used for search: -H2O; y; -NH3; y; b; b; -H2O; b; -NH3; y

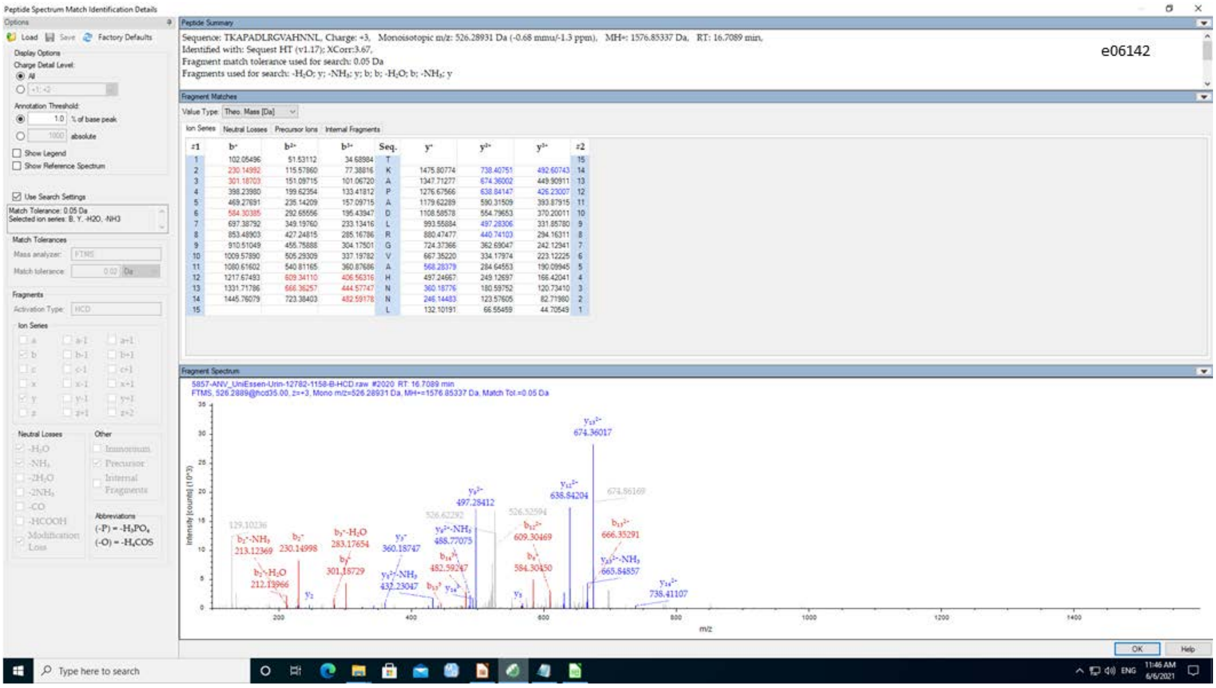
e06142
Sequence: TKAPADLRGVAHNNL, Charge: +3, Monoisotopic m/z: 526.28931 Da (-0.68 mmu/-1.3 ppm), MH+: 1576.85337 Da, RT: 16.7089 min,
Identified with: Sequest HT (v1.17); XCorr:3.67,
Fragment match tolerance used for search: 0.05 Da
Fragments used for search: -H2O; y; -NH3; y; b; b; -H2O; b; -NH3; y

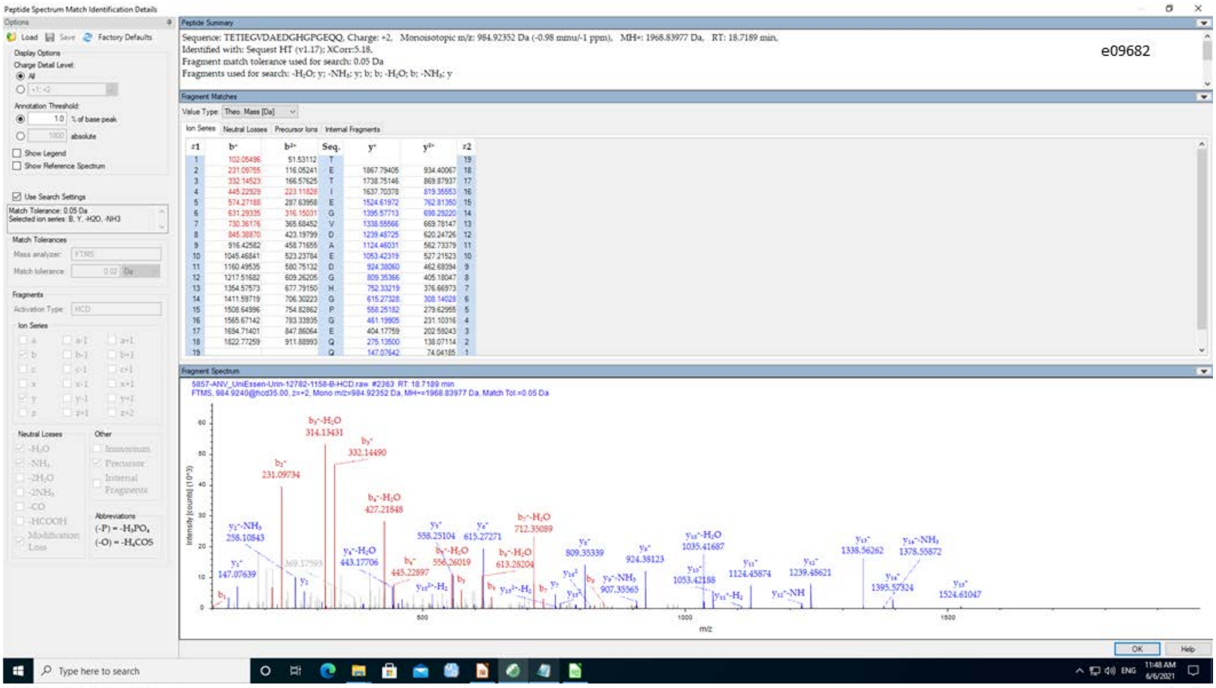
Peptide Spectrum Match Identification Details
e09682
Sequence: TETIEGVDAEDGHGPGEQQ, Charge: +2, Monoisotopic m/z: 984.92352 Da (-0.98 mmu/-1 ppm), MH+: 1968.83977 Da, RT: 18.7189 min,
Identified with: Sequest HT (v1.17); XCorr:5.18,
Fragment match tolerance used for search: 0.05 Da
Fragments used for search: -H2O; y; -NH3; y; b; b; -H2O; b; -NH3; y
5857-ANV_UniEssen-Urin-12782-1158-B-HCD.raw #2363 RT: 18.7189 min
FTMS, 984.9240@hcd35.00, z=+2, Mono m/z=984.92352 Da, MH+=1968.83977 Da, Match Tol.=0.05 Da
Intensity [counts] (10^3)
m/z

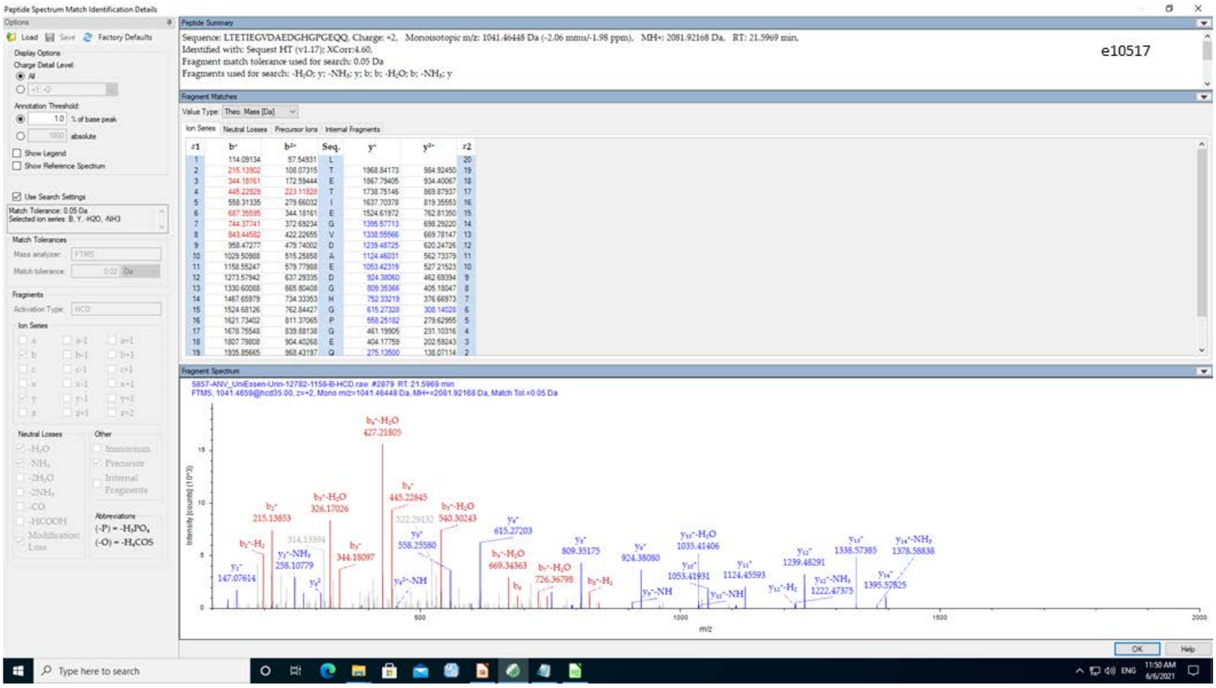
Peptide Spectrum Match Identification Details
e10517
Sequence: LTETIEGVDAEDGHGPGEQQ, Charge: +2, Monoisotopic m/z: 1041.46448 Da (-2.06 mmu/-1.98 ppm), MH+: 2081.92168 Da, RT: 21.5969 min,
Identified with: Sequest HT (v1.17); XCorr:4.60,
Fragment match tolerance used for search: 0.05 Da
Fragments used for search: -H2O; y; -NH3; y; b; b; -H2O; b; -NH3; y
5857-ANV_UniEssen-Urin-12782-1158-B-HCD.raw #2879 RT: 21.5969 min
FTMS, 1041.4659@hcd35.00, z=+2, Mono m/z=1041.46448 Da, MH+=2081.92168 Da, Match Tol.=0.05 Da
Intensity [counts] (10^3)
m/z

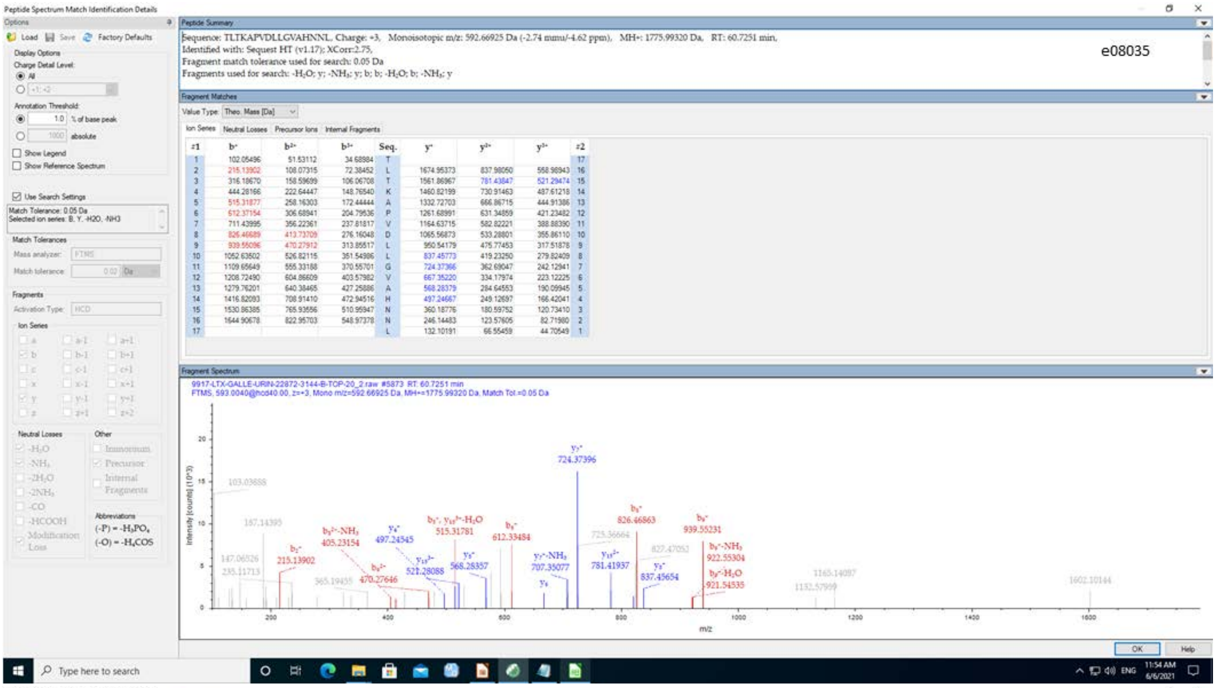

Peptide Spectrum Match Identification Details
e08035
Sequence: TLTKAPVDLLGVAHNNL, Charge: +3, Monoisotopic m/z: 592.66925 Da (-2.74 mmu/-4.62 ppm), MH+: 1775.99320 Da, RT: 60.7251 min,
Identified with: Sequest HT (v1.17); XCorr:2.75,
Fragment match tolerance used for search: 0.05 Da
Fragments used for search: -H2O; y; -NH3; y; b; b; -H2O; b; -NH3; y


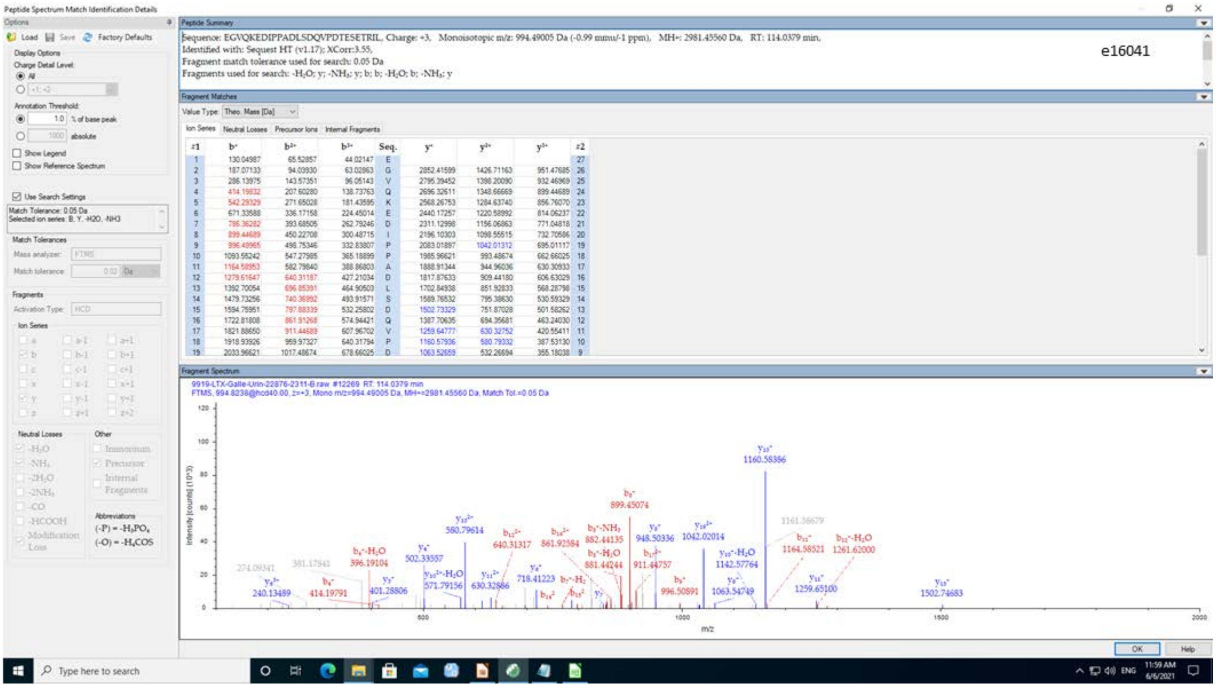

Peptide Spectrum Match Identification Details
e16041
Sequence: EGVQKEDIPPADLSDQVPDTESETRIL, Charge: +3, Monoisotopic m/z: 994.49005 Da (-0.99 mmu/-1 ppm), MH+: 2981.45560 Da, RT: 114.0379 min,
Identified with: Sequest HT (v1.17); XCorr:3.55,
Fragment match tolerance used for search: 0.05 Da
Fragments used for search: -H2O; y; -NH3; y; b; b; -H2O; b; -NH3; y

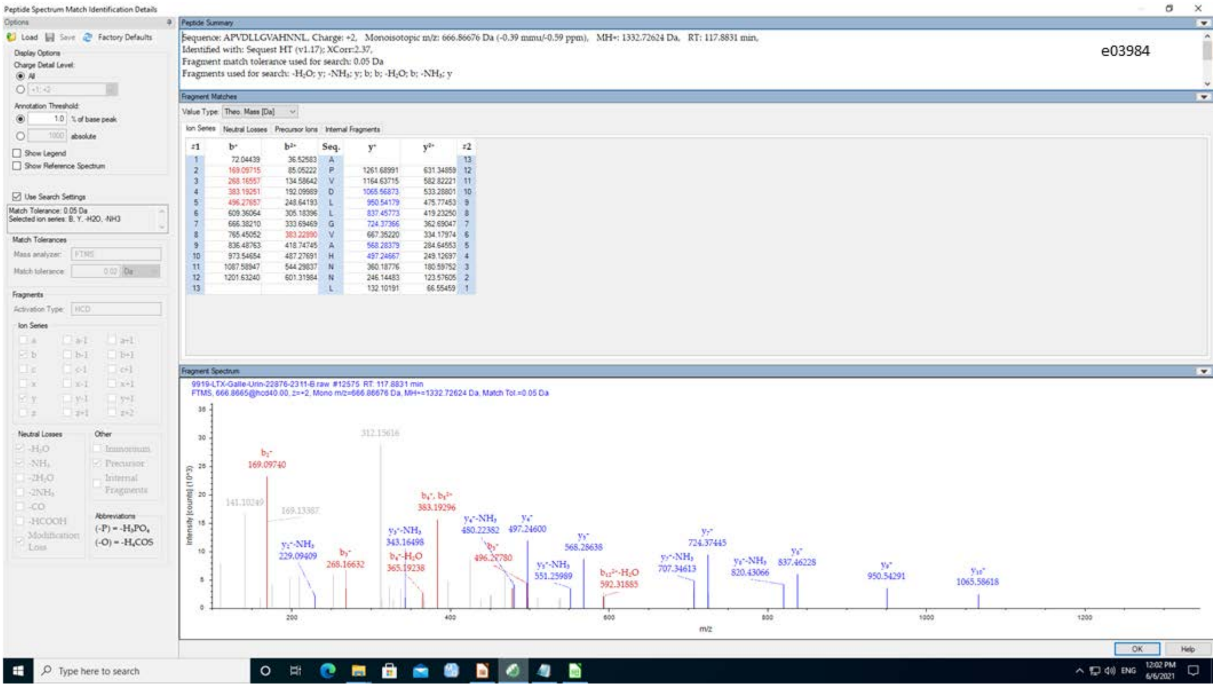


**Supplementary Figure 1:** Tandem mass spectra and assignment of sequences of the 23 complement derived urine peptides detected.

**Supplementary Figure 2**

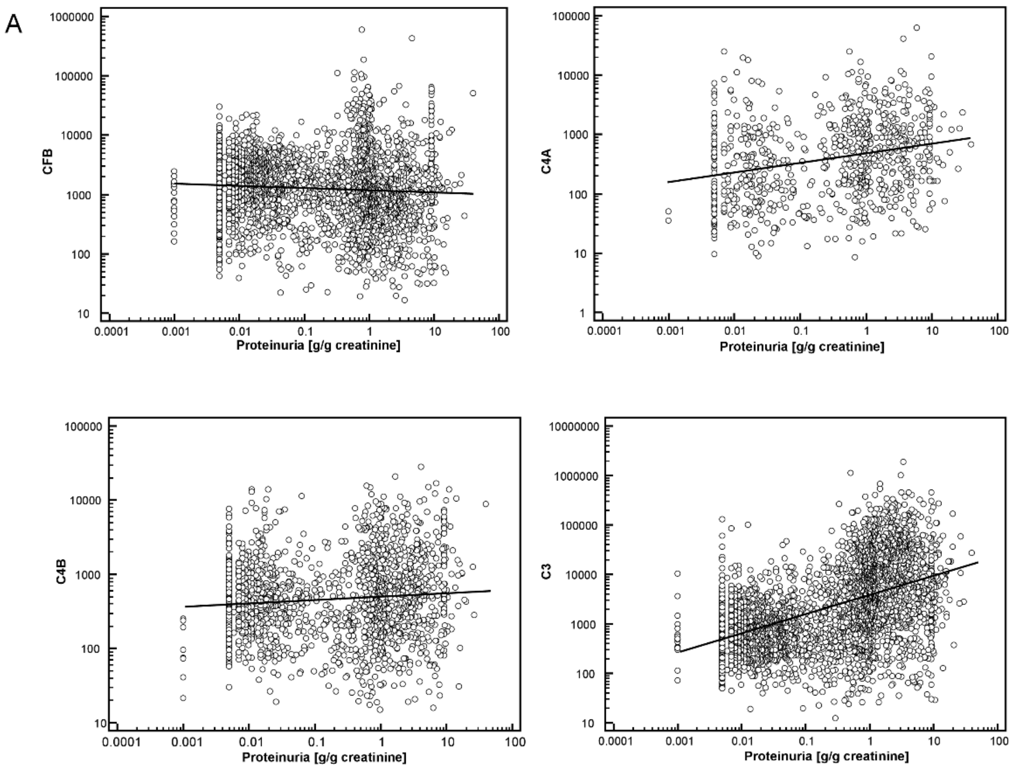


**Supplementary Figure 2:** Association of the combined abundance of complement peptides (from Complement factor B, 4A, 4B and 3) with proteinuria, A) prior and B) after adjustment for proteinuria.

**Supplementary Figure 3**

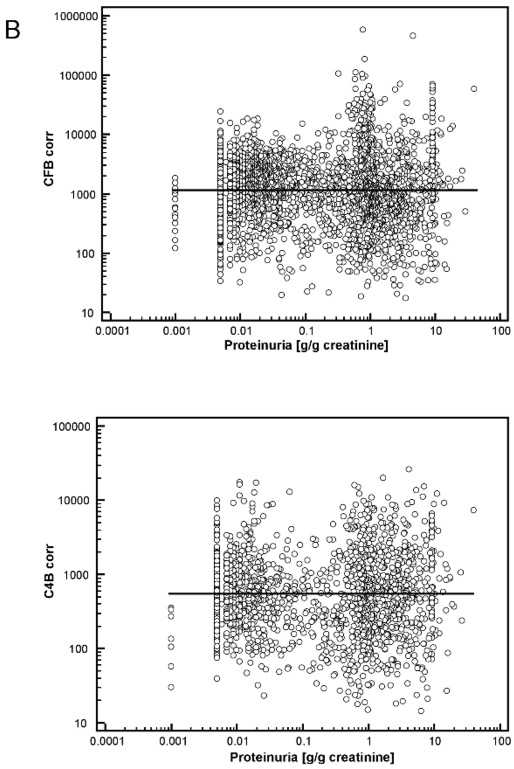


**Supplementary Figure 3:** Association of proteinuria with eGFR in the study population. As expected, a highly significant correlation is detectable.

| N | disease/condition | e13403 | e11594 | e11703 | e12606 | e10517 | e08566 | e09682 | e08161 | e09265 | e06142 | e08035 | e03984 | e00117 | e09429 | e16041 | e17084 | e18666 | e12939 | e14381 | e16109 | e06787 | e08849 | e09686 |
|---|---|---|---|---|---|---|---|---|---|---|---|---|---|---|---|---|---|---|---|---|---|---|---|---|
| 422 | acute_kidney_injury | 56,01 | 2,94 | 0,47 | 31,43 | 44,29 | 0,83 | 0,33 | 22,63 | 1,15 | 18,90 | 45,07 | 1,16 | 2,65 | 55,02 | 5,38 | 0,58 | 0,29 | 57,80 | 21,84 | 27,08 | 1,04 | 11,90 | 7,81 |
| 273 | ADPKD | 7,13 | 3,81 | 3,66 | 1,62 | 0,80 | 1,96 | 1,23 | 0,66 | 2,16 | 0,35 | 0,26 | 0,20 | 0,57 | 5,34 | 0,82 | 0,97 | 0,90 | 5,74 | 1,48 | 0,37 | 1,39 | 1,15 | 2,20 |
| 8 | amyloidosis | 0,00 | 2,89 | 0,29 | 1,43 | 2,96 | 0,26 | 0,36 | 24,43 | 0,36 | 0,00 | 0,00 | 0,00 | 0,00 | 0,00 | 1,51 | 0,06 | 0,00 | 38,01 | 1,15 | 0,00 | 2,33 | 0,00 | 8,90 |
| 8 | atypical_hemolytic_uremic_syndrome | 0,00 | 0,07 | 0,00 | 0,44 | 0,00 | 0,89 | 0,00 | 0,00 | 0,16 | 1,15 | 0,00 | 0,00 | 0,00 | 33,93 | 0,66 | 0,15 | 0,00 | 0,33 | 0,78 | 0,00 | 0,15 | 0,00 | 0,00 |
| 24 | C3_glomerulopathy | 0,00 | 0,31 | 0,31 | 0,49 | 4,72 | 0,50 | 3,68 | 5,44 | 0,83 | 5,18 | 14,68 | 0,00 | 2,04 | 293,83 | 0,55 | 0,00 | 0,00 | 60,96 | 7,93 | 0,00 | 1,37 | 5,49 | 2,03 |
| 567 | CAKUT | 14,23 | 6,50 | 0,99 | 1,70 | 1,00 | 0,40 | 13,18 | 0,61 | 3,01 | 0,15 | 2,69 | 0,46 | 0,46 | 0,23 | 0,34 | 1,61 | 3,87 | 4,72 | 1,89 | 23,21 | 0,40 | 1,39 | 2,15 |
| 4428 | DiabetesMell_yes | 0,31 | 1,72 | 0,51 | 1,59 | 9,58 | 1,43 | 0,77 | 3,31 | 1,82 | 1,42 | 9,11 | 0,75 | 1,75 | 6,69 | 1,76 | 0,71 | 0,84 | 4,54 | 1,37 | 2,23 | 0,96 | 3,01 | 1,28 |
| 1401 | diabetic_nephropathy | 2,57 | 0,83 | 0,18 | 1,79 | 3,35 | 0,62 | 0,20 | 39,53 | 0,84 | 6,26 | 10,74 | 5,70 | 0,97 | 179,37 | 2,23 | 0,12 | 0,36 | 129,57 | 8,21 | 2,98 | 2,30 | 4,68 | 5,78 |
| 12 | fanconi_syndrom | 3,46 | 2,23 | 0,07 | 0,95 | 0,00 | 0,00 | 4,15 | 0,00 | 1,99 | 0,21 | 0,00 | 0,00 | 0,00 | 0,00 | 2,10 | 0,00 | 0,41 | 0,15 | 5,15 | 1,80 | 0,61 | 9,37 | 0,00 |
| 126 | FSGS | 2,42 | 1,83 | 0,35 | 5,61 | 4,26 | 0,81 | 1,54 | 25,26 | 1,75 | 7,19 | 64,36 | 1,91 | 3,76 | 50,13 | 3,40 | 0,20 | 0,51 | 206,63 | 5,96 | 150,08 | 1,43 | 4,62 | 9,23 |
| 811 | IgAN | 20,24 | 1,68 | 0,56 | 1,59 | 6,29 | 0,78 | 0,75 | 10,81 | 1,11 | 5,16 | 65,72 | 0,56 | 3,11 | 109,19 | 1,67 | 0,30 | 0,09 | 114,80 | 10,21 | 7,02 | 2,40 | 4,70 | 127,85 |
| 50 | MCD | 0,00 | 9,41 | 0,82 | 0,50 | 8,54 | 2,08 | 0,45 | 31,91 | 1,19 | 2,18 | 0,00 | 0,00 | 1,47 | 358,95 | 0,32 | 0,07 | 0,00 | 452,90 | 23,43 | 79,50 | 4,10 | 12,80 | 0,94 |
| 113 | MGN | 0,86 | 1,68 | 0,22 | 2,72 | 8,56 | 0,96 | 0,61 | 111,32 | 2,05 | 15,72 | 26,65 | 0,36 | 3,13 | 79,38 | 3,01 | 0,04 | 0,00 | 176,52 | 13,61 | 0,36 | 4,34 | 26,57 | 20,77 |
| 66 | Morbus_Fabry | 12,18 | 2,66 | 0,10 | 1,02 | 4,02 | 0,86 | 1,99 | 1,42 | 1,11 | 1,33 | 0,00 | 0,67 | 0,52 | 30,88 | 0,54 | 0,85 | 0,26 | 12,56 | 2,94 | 1,27 | 0,73 | 8,63 | 2,04 |
| 50 | MPGN | 33,52 | 1,86 | 2,19 | 12,69 | 11,41 | 0,94 | 22,91 | 16,14 | 1,61 | 7,13 | 38,93 | 0,24 | 5,13 | 56,08 | 2,85 | 0,04 | 0,21 | 82,29 | 6,61 | 3,56 | 2,13 | 4,43 | 3,20 |
| 104 | hypertensive nephrosclerosis | 5,89 | 1,68 | 0,11 | 5,37 | 34,01 | 0,43 | 0,24 | 1,36 | 0,61 | 2,11 | 6,58 | 0,00 | 1,75 | 25,74 | 13,37 | 0,33 | 0,92 | 35,43 | 2,77 | 0,16 | 0,73 | 22,64 | 4,10 |
| 127 | nephrotic_syndrom | 45,54 | 2,66 | 2,58 | 1,39 | 5,90 | 2,42 | 15,89 | 104,96 | 7,52 | 21,36 | 33,29 | 4,40 | 3,10 | 353,70 | 0,84 | 0,94 | 0,33 | 256,27 | 20,75 | 120,66 | 2,69 | 211,49 | 2,70 |
| 2300 | NTx | 17,16 | 0,94 | 0,26 | 3,40 | 1,87 | 0,50 | 1,02 | 5,60 | 0,71 | 3,63 | 32,61 | 0,77 | 3,33 | 8,45 | 1,38 | 0,67 | 0,21 | 11,72 | 3,75 | 2,87 | 1,40 | 7,59 | 2,91 |
| 20 | SLE | 0,00 | 0,04 | 0,00 | 0,99 | 0,00 | 0,34 | 0,00 | 0,00 | 0,48 | 0,00 | 0,00 | 0,00 | 0,11 | 0,00 | 1,81 | 0,08 | 4,83 | 0,00 | 0,56 | 0,00 | 0,33 | 0,00 | 0,00 |
| 94 | SLE_nephritis | 1,65 | 1,44 | 1,18 | 2,06 | 6,63 | 0,48 | 0,21 | 9,48 | 1,51 | 3,96 | 54,33 | 4,72 | 0,52 | 71,33 | 3,01 | 0,19 | 3,73 | 131,44 | 5,07 | 1,17 | 1,10 | 7,32 | 7,77 |
| 159 | vaskulitis | 31,61 | 0,47 | 0,05 | 16,78 | 6,82 | 0,33 | 0,23 | 2,89 | 0,25 | 1,47 | 54,30 | 0,04 | 7,60 | 33,40 | 6,93 | 0,25 | 1,37 | 36,48 | 2,97 | 6,88 | 0,68 | 3,13 | 9,00 |
| 4864 | normal_control | 1,00 | 1,00 | 1,00 | 1,00 | 1,00 | 1,00 | 1,00 | 1,00 | 1,00 | 1,00 | 1,00 | 1,00 | 1,00 | 1,00 | 1,00 | 1,00 | 1,00 | 1,00 | 1,00 | 1,00 | 1,00 | 1,00 | 1,00 |

**Supplementary Table 1** Relative abundance of all 23 collagen derived peptides, listed per disease group, normalized to the normal controls.
